# A Framework for Variant Filtering to Improve Diagnostic Outcomes in Genome Sequencing

**DOI:** 10.64898/2026.11.07.26367371

**Authors:** Rivka Birnbaum, Kamila Lee, Yaron Mazor, Yaara Unger, Shira Yanovsky-Dagan, Itay Grundwag, Michal Chalk, Nava Shaul-Lotan, Lior Greenbaum, Orit Freireich, Tehila Klopstock, Tal Dror, Meyrav Sebbagh, Rachel Beeri, Omer Murik, Dan Raved, Rachel Rock, Ben Pode-Shakked, Hagit Shani, Sagi Josefsberg, Tamar Harel, Hagar Mor-Shaked

**Author notes:** Corresponding author: Hagar Mor-Shaked, Department of Genetics, Hadassah-Hebrew University Medical Center, POB 12000, Jerusalem, Israel. These authors contributed equally (Rivka Birnbaum, Kamila Lee).

## Abstract

Rare genetic disorders frequently remain undiagnosed despite the use of exome sequencing (ES) and genome sequencing (GS). This study aimed to enhance diagnostic yield by introducing a systematic framework: SMART filtering (Systematic, Multi-step, Automated, Rare variant prioritization Tool), designed to optimize GS analysis, with a particular emphasis on non-coding regions. We retrospectively analyzed 153 cases with previously negative ES and GS results. SMART filtering was applied as a pre-annotation filtering step, integrating conservation metrics, allele frequency thresholds, and splicing prediction scores. Then followed by phenotype-based prioritization and manual review. Application of this framework identified pathogenic or likely pathogenic variants in 4.6% of cases, strong variants of uncertain significance (VUS) in 1.3%, and additional candidate variants in 5.2% of cases. Several deep-intronic variants were confirmed by RNA studies and segregation analyses. The framework enabled efficient prioritization of non-coding and regulatory variants that were overlooked by standard analyses.

In conclusion, SMART filtering offers a reproducible and scalable strategy to improve diagnostic yield in GS, particularly in cases where conventional analysis methods fall short. Integrating such frameworks into clinical genomic pipelines can enhance variant interpretability and diagnostic performance, ultimately leading to better patient care outcomes.

## INTRODUCTION

Genetic testing for rare diseases, particularly exome sequencing (ES), achieves a diagnostic yield of 25–35% (Alfares et al. 2018). This yield is notably higher in family trios and consanguineous populations (Clark et al. 2018; Alfares et al. 2018). Such testing plays a critical role in facilitating early diagnosis, enabling targeted therapeutic interventions, and supporting informed reproductive decision-making.

ES is considered a first-tier test in the genetic diagnostic pipeline (Boycott et al. 2017; Srivastava et al. 2019; Marwaha et al. 2022), but when ES results are unrevealing, it is recommended to proceed to more comprehensive technologies, including genome sequencing (GS) (Bagnall et al. 2018; Rajagopalan et al. 2021; Marwaha et al. 2022). GS captures approximately 98% of the human genome, including intronic variants, complex structural variants, and protein-coding variants, which may not be accurately captured or may be impossible to detect using ES (Marwaha et al. 2022). When employed as the first and sole diagnostic tool, GS has the potential to achieve a diagnostic rate of up to 43% (Clark et al. 2018; Manickam et al. 2021). Accordingly, the diagnostic yield using GS following a negative ES can be improved by ∼10% (Alfares et al. 2018; Bertoli-Avella et al. 2021; Wojcik et al. 2024; Albuquerque et al. 2025), mainly depending on the clinical presentation.

The main challenges of analyzing GS data include considerable, although rapidly declining, costs (Nisar et al. 2021), and absence of comprehensive data for non-coding region annotation (Boycott et al. 2017; Kernohan and Boycott 2024). This challenge arises from the limited availability of publicly accessible whole genome databases of healthy individuals, which are essential for effective comparison, and leaves a massive number of variants for manual analysis.

Additionally, there is a significant under-reporting of variants in non-coding regions (Ellingford et al. 2022), including those known to regulate well-established disease-causing genes. Another challenge is the lack of guidance and standards for genomic analysis and its clinical implementation, and those available for genetic analysis, do not extend to detailed instructions for non-coding regions (Boycott et al. 2017; Marshall et al. 2020). Recently published recommendations address the interpretation of intronic variants and the distinct approach required to establish and report a diagnosis (Ellingford et al. 2022); however, these are not yet considered consensus guidelines.

Due to these challenges and the fact that most published GS-based diagnoses originate from exonic regions, genomes are often analyzed with an exome-focused approach, leveraging the superior capture performance of GS in those areas (Umlai et al. 2022). While analyzing non-exonic regions is expected to increase diagnostic yield, there is no consensus framework for non-coding variant filtering.

With the increasing use of GS, vast amounts of data are generated, including a massive number of variants (Pena and Tarazona-Santos 2022; Bagger et al. 2024), which require comprehensive filtering steps to identify the “culprit” variant responsible for the disease (Pedersen et al. 2021). Over the years, various general filtering strategies have been proposed to streamline analysis (Ravasio et al. 2018). Common approaches include phenotype-based filtering using human phenotype ontology (HPO) terminology, genotype-phenotype associations, and bioinformatic predictions such as tolerance to loss-of-function variants and conservation metrics (e.g., CADD (Moore et al. 2023), GERP (Huber et al. 2020), SpliceAI (Jaganathan et al. 2019), and AlphaMissense (Cheng et al. 2023).

In this work, we present a novel framework for efficient filtering of genomic data, followed by a systematic genomic analysis for the diagnosis of rare genetic diseases in previously negative cases. This approach, which we named SMART filtering (Systematic Multi-step Automated Rare variant prioritization Tool), allows a genome-wide analysis, while eliminating common variants and variants with low bioinformatic predictions. Combined with phenotype correlation, SMART assists in identifying non-coding candidates that might otherwise be overlooked.

## MATERIALS AND METHODS

### Participants

We evaluated 153 patients with diverse phenotypes referred for GS in Hadassah Medical Center between 2021 and 2025. All patients had previously negative results from ES, chromosomal microarray analysis (CMA), and clinical GS analyses. Inclusion required a high clinical suspicion for a monogenic disorder, based on a phenotype suggestive of a Mendelian disorder, consanguinity, or family history. Cases in which the primary indication suggested a multifactorial etiology were excluded. All participants provided informed consent, and clinical information supplied by referring clinicians was incorporated into the analysis. The study was conducted in accordance with IRB-approved protocol 0306-10-HMO. All 153 genomes in this study were analyzed as singleton cases.

### Genome sequencing

Genomic DNA was extracted from peripheral blood and processed using the Illumina PCR-free TruSeq DNA Library Prep Kit. Sequences were generated on an Illumina NovaSeq 6000 platform (150 bp paired end reads; minimum depth of 30X coverage). FASTQ files were analyzed on the Geneyx Analysis platform (Dahary et al. 2019). Alignment and calling of single nucleotide variations (SNVs) and copy number variants (CNVs) were performed using the Illumina DRAGEN Bio-IT pipeline, followed by comprehensive annotation. Samples were aligned to GRCh37/hg19 (118 samples) or GRCh38/hg38 (35 samples) according to clinical availability.

### SMART filtering

SMART filtering was implemented as an automated pre-annotation filtering step. Variants were restricted to an ‘all-genes’ BED file comprising the full span of all RefSeq transcripts (ncbiRefSeq.txt.gz file) with ±5 kb padding to incorporate regulatory regions. Pre-annotation filtering thresholds included CADD ≥5, SpliceAI ≥0.049, and gnomAD v3.1 allele count of 0 for homozygous/hemizygous variants and ≤300 for heterozygous variants, based on empirical evaluation and recommendations for intronic variant interpretation (Barbosa et al. 2022). BED files for GRCh37 and GRCh38 are available upon request.

The ‘all-genes’ BED file spans 2.74 Gb in GRCh38 and 2.29 Gb in GRCh37. Filtering efficiency was assessed by recording variant counts at each step in four representative genomes (three GRCh38, one GRCh37).

### Variant annotation

Following SMART filtering, retained variants were annotated using 34 and 38 datasets for GRCh37 and GRCh38, respectively, integrated within the Geneyx platform. The full list of data sources and versions is provided in the Supplementary Methods under the “Variant annotation sources” section.

### Phenotyper for phenotype prioritization

Phenotype-driven gene prioritization was performed using an internally developed Geneyx algorithm that queries a harmonized knowledge graph built from approximately 50 databases originated in 29 curated sources (including OMIM, HPO, ClinVar, Orphanet, UniProt, and LitVar2). Patient HPO terms, derived from the referral documents following genetic counseling, were matched against gene–phenotype associations to generate a composite prioritization score; with scores ≥20 indicating robust correlation. Candidate genes were flagged when their score was ≥20 or when they matched most of the patient’s HPO terms. Databases were updated every 3-4 months with validation for schema and index integrity, and pipelines were version-controlled. A full description of the Phenotyper, scoring framework and data sources is provided in the Supplementary Methods.

### Filtering and systematic variant review

Annotations and Phenotyper scores were reviewed independently by two analysts. Discrepancies between analysts were resolved by consensus.

Preset-automatic filters for rare variant identification required: for AR disorders <2 homozygotes in the local cohort and none in gnomAD; for AD disorders, <4 heterozygotes locally, <2 in gnomAD and no hemizygotes or homozygotes in both databases. All variants required a Phenotyper score ≥20. Variants without CADD or SpliceAI scores were also kept when absent from both databases if they had a Phenotyper score >150. (Fig 1A) Potential compound heterozygous variants were evaluated using criteria similar to those applied to homozygous variants, with emphasis on rarity and predicted functional impact.

**Figure 1.**
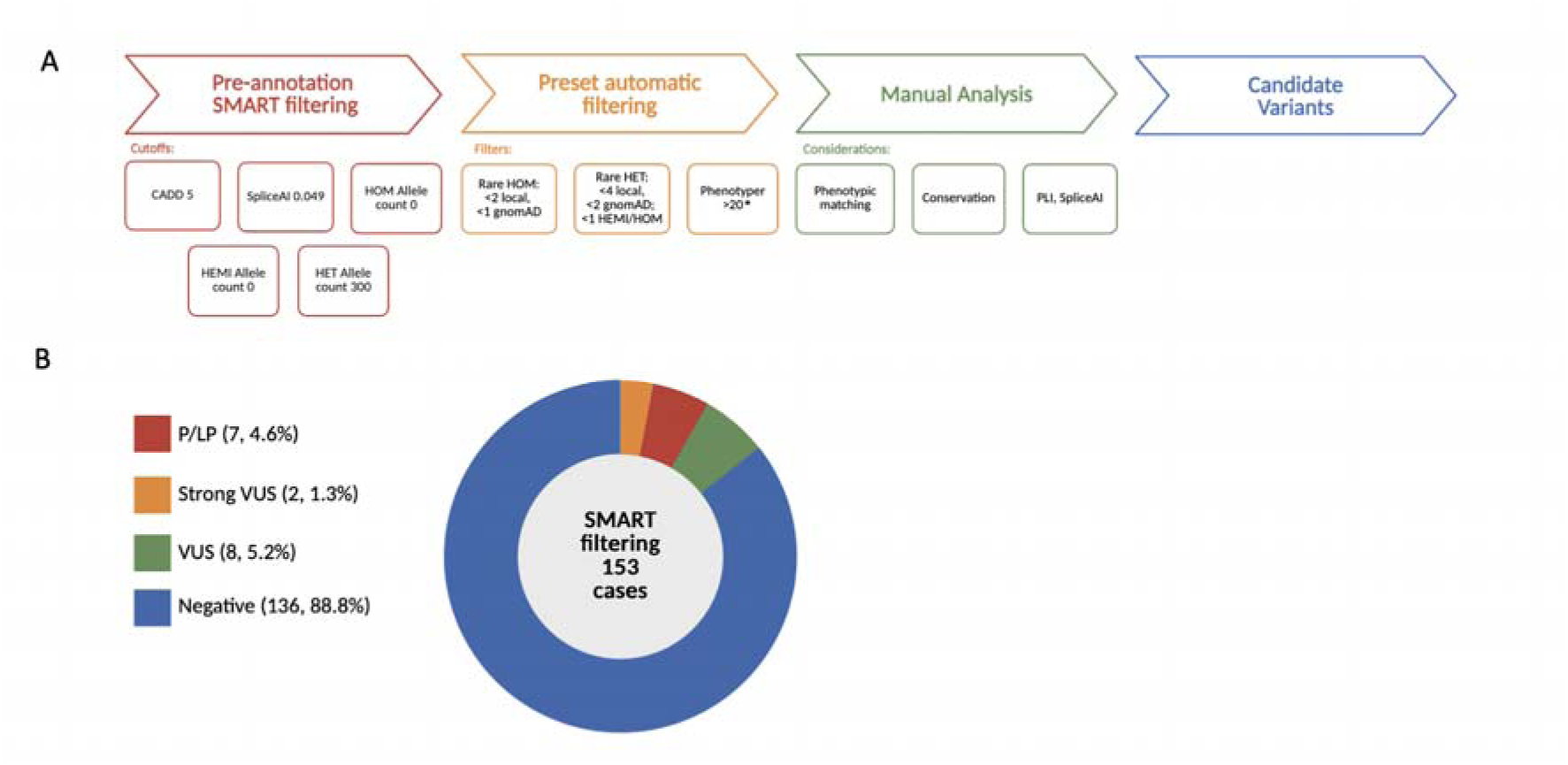
Genetic analysis process and inclusion criteria for case interpretation. (A) Overview of the SMART filtering process. The pipeline consists of three major stages leading to the identification of candidate variants: pre-annotation SMART filtering, preset automatic filtering and manual analysis. The resulting candidate variants are reviewed for clinical relevance. *Variants lacking CADD or SpliceAI scores that were absent from gnomAD and the local cohort were retained if Phenotyper >150. (B) The donut chart illustrates the diagnostic outcomes of SMART filtering in a cohort of 153 cases. The final classification of variants following filtering and manual review. P/LP: 7 cases (4.6%), Strong VUS, yet to be validated: 2 cases (1.3%), VUS: 8 cases (5.2%), Negative: 136 cases (88.8%).

Following preset automatic filtering, candidates were sorted by Phenotyper score and reviewed in descending rank order. The review proceeded gene-wise rather than variant-wise: variants were grouped by gene, and genes lacking an established or plausible disease association consistent with the proband’s phenotype were de-prioritized as a group, reducing the effective number of interpretive units per case.

Within the retained list, prioritization focused on compatibility with the expected inheritance model and zygosity, rarity in gnomAD and the local cohort, ClinVar and LitVar2 evidence, gene constraint (probability of loss-of-function intolerance, pLI), conservation metrics (GERP-RS, CADD) and predicted splicing impact (SpliceAI). Conservation and predictive metrics were considered qualitatively without additional fixed thresholds.

Variant rarity was evaluated with gnomAD v3.1 and a local cohort as a reference, comprising 26,593 exomes and 497 genomes from Hadassah Medical Center, using zygosity-specific thresholds.

Intronic variants located more than 100 bp from exon boundaries were considered deep-intronic (Vaz-Drago et al. 2017).

Manual interpretation used additional population resources, including dbSNP (Sherry et al. 2001; Phan et al. 2025) and CoLoRSdb (Lake and Consortium of Long Read Sequencing (CoLoRS) 2024) to flag known common variants.

### Ranking performance assessment

To quantify the prioritization efficiency of the SMART workflow (‘all-genes’ BED with pre-annotation filter) versus the default (‘exons-only’ BED, no pre-annotation filter) approach, we recorded the rank position of all reported and VUS variants within the candidate list generated after preset automatic filtering in both pipelines. In cases with two pathogenic variants (compound heterozygous or digenic), both variants were ranked independently. To enable comparison across cases with differing candidate pool sizes, we calculated a normalized rank for each variant as:

Normalized rank = Rank position / Total number of candidates after filtering

The normalized rank is a dimensionless value in the range [0, 1], where values approaching 0 indicate that the diagnostic variant was placed near the top of the prioritized list relative to the total candidate burden. Median normalized rank was computed across all diagnostic probands, using both variants if two variants existed per gene. Comparisons were made between the default and SMART conditions to assess the added value of the SMART pre-annotation filtering step.

### RNA Sequencing

RNA-seq libraries were prepared using the KAPPA RNA HyperPrep kit and paired-end sequencing was performed on an Illumina NovaSeq 6000 platform. The resulting FASTQ files (transferable upon request) were computationally processed in-house, using SAMtools Mappings Indexer (v2.1.1) and Star_align (v0.0.1). RNA-seq data were aligned to the GRCh37/hg19 reference genome and lifted over to GRCh38 for specific coordinates of interest. Analysis was performed by focused visualization, using IGV (v2.6.6) and compared with an internal normal dataset (from the same tissue).

### RNA studies

Targeted RNA studies for selected genes (*PAX3*, *SLC26A4*) were performed using reverse-transcriptase polymerase chain reaction (RT-PCR) and Sanger sequencing. RNA was extracted from fibroblasts for *PAX3* analysis or using the Tempus™ Spin RNA Isolation Kit for the *SLC26A4* analysis. Primer sequences and PCR conditions are provided in the Supplementary Methods. PCR products were examined by agarose gel electrophoresis and sequenced to assess aberrant splicing.

### RNA-seq data processing and analysis

RNA-seq data were processed using the OmicsDiscoveries RNAvision™ platform (v1.1.0). Alignment was performed using the nf-core/rnaseq pipeline (v3.19.0) with the aligner = star_salmon option, including read trimming (Trim Galore v0.6.10) and STAR alignment (v2.7.11b; - -twopassMode = BASIC). Quality control was performed with Qualimap (v2.3), RSeQC (v5.0.2) (Wang et al. 2012), SAMtools (v1.21), and MultiQC (v1.29.0). Sample identity, sex concordance, and tissue verification were assessed with DROP v1.5.0 (Yépez et al. 2021).

Aberrant expression was detected using the DROP aberrant expression module with GENCODE v48 (GRCh38). Read counting used summarizeOverlaps from the GenomicAlignments (Lawrence et al. 2013) R package, and genes with a 95th-percentile FPKM < 0.5 were excluded. Expression outliers were identified with OUTRIDER v1.26.3 (Brechtmann et al. 2018). Aberrant splicing was detected with DROP based on FRASER v2.4.6 (Scheller et al. 2023), using the intronic Jaccard index. Mono-allelic expression used GATK (v4.4) ASEReadCounter function (Castel et al. 2015), excluding sites with <10 total reads.

To increase statistical power, 95 additional matched samples were jointly processed and included in model fitting. Aberrant events were visually inspected using RNAvision™ and IGV. A full description is provided in the Supplementary Methods.

### Cell culture and Cycloheximide (CHX) treatment

Skin fibroblasts were cultured under standard conditions (DMEM, 15% fetal bovine serum, penicillin–streptomycin, L-glutamine; 37°C; 5% CO₂; Biological Industries Beit Haemek, Israel). For nonsense-mediated decay (NMD) inhibition, cells were treated with cycloheximide (0.5 mg/mL; Cat. No. 239765 Sigma-Aldrich) for 24 hours prior to RNA extraction.

## RESULTS

### SMART filtering analysis

Across four representative genomes, each containing approximately 4.9 million SNVs, restriction to the ‘all-genes’ BED file retained 70–84% of variants (3.4–4.2 million per genome), reflecting intentionally permissive pre-annotation thresholds. The subsequent pre-annotation SMART filtering step removed a further ∼98%, leaving 55,549-87,193 candidates per genome (<2% of the original variant set; Table S1). The filtered VCF proceeded to full annotation, phenotype correlation, preset automatic filtering and manual review (Fig 1).

We re-analyzed 153 previously undiagnosed cases (negative by ES and clinical GS) using SMART filtering. Nine cases (5.9%) carried reportable variants: seven pathogenic (P) or likely pathogenic (LP) (4.6%) and two novel highly relevant variants of uncertain significance (Strong VUS), pending validation (1.3%). Eight additional cases (5.2%) harbored lower-certainty VUS (Fig 1, Table 1, Table S2). Cases were categorized as having reported variants, VUS or negative, according to integrated clinical and molecular evidence. The reportable variant group comprised nine cases: seven harbored intronic variants classified as P/LP according to ACMG criteria, of them, three were deep-intronic variants. These variants showed strong phenotypic concordance, defined by a Phenotyper score of ≥20 or concordance with most HPO terms, with supportive bioinformatic data such as rarity, conservation and in silico prediction scores. Where available, segregation analysis, RNA studies, and published evidence further supported classification. Two additional cases harbored strong VUS suggestive of a likely, but not yet definitive, diagnosis. The VUS group included eight variants with partial phenotype overlap or genes with limited evidence, for which current data were insufficient to justify further workup (Table S2).

**Table 1.**
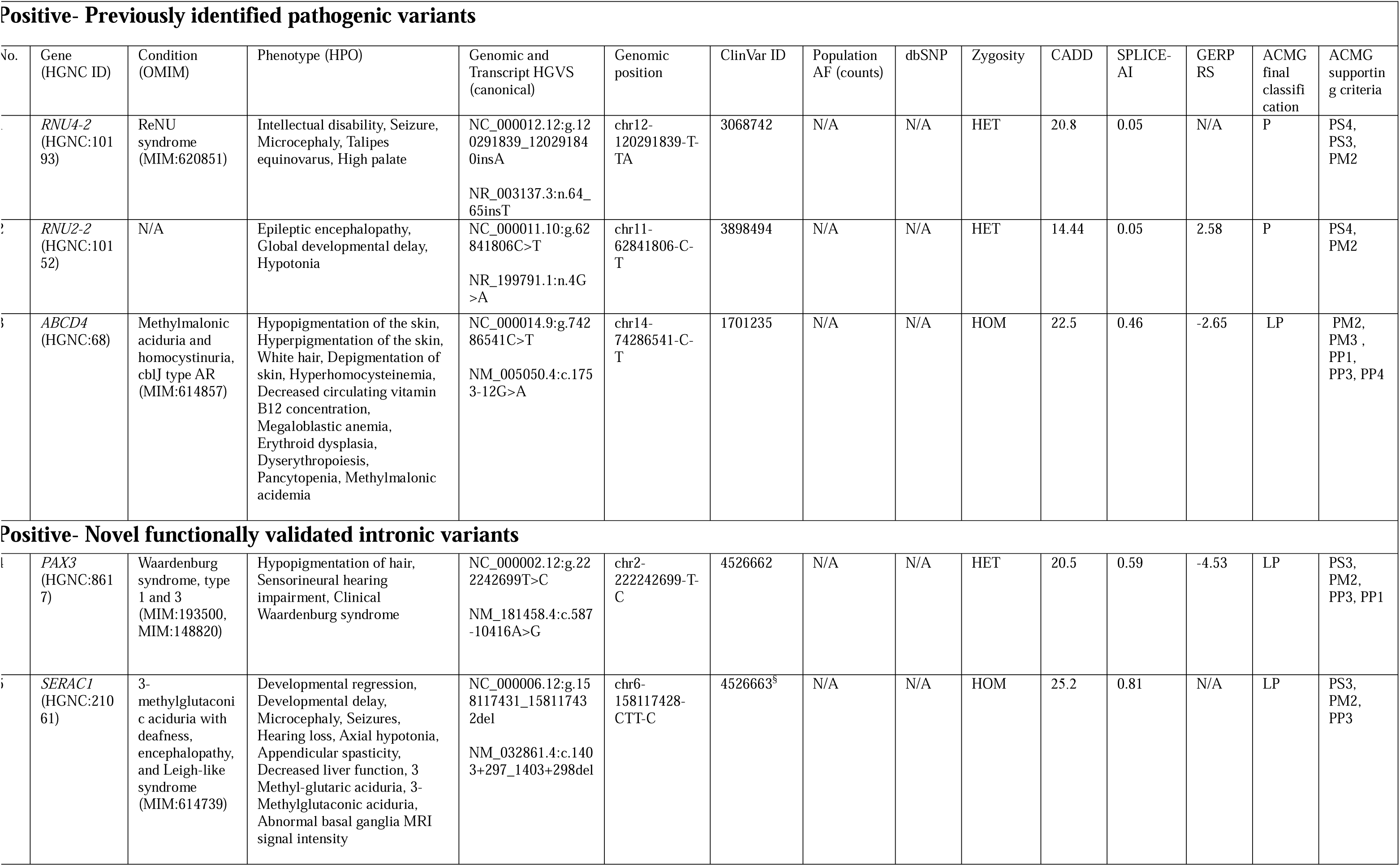
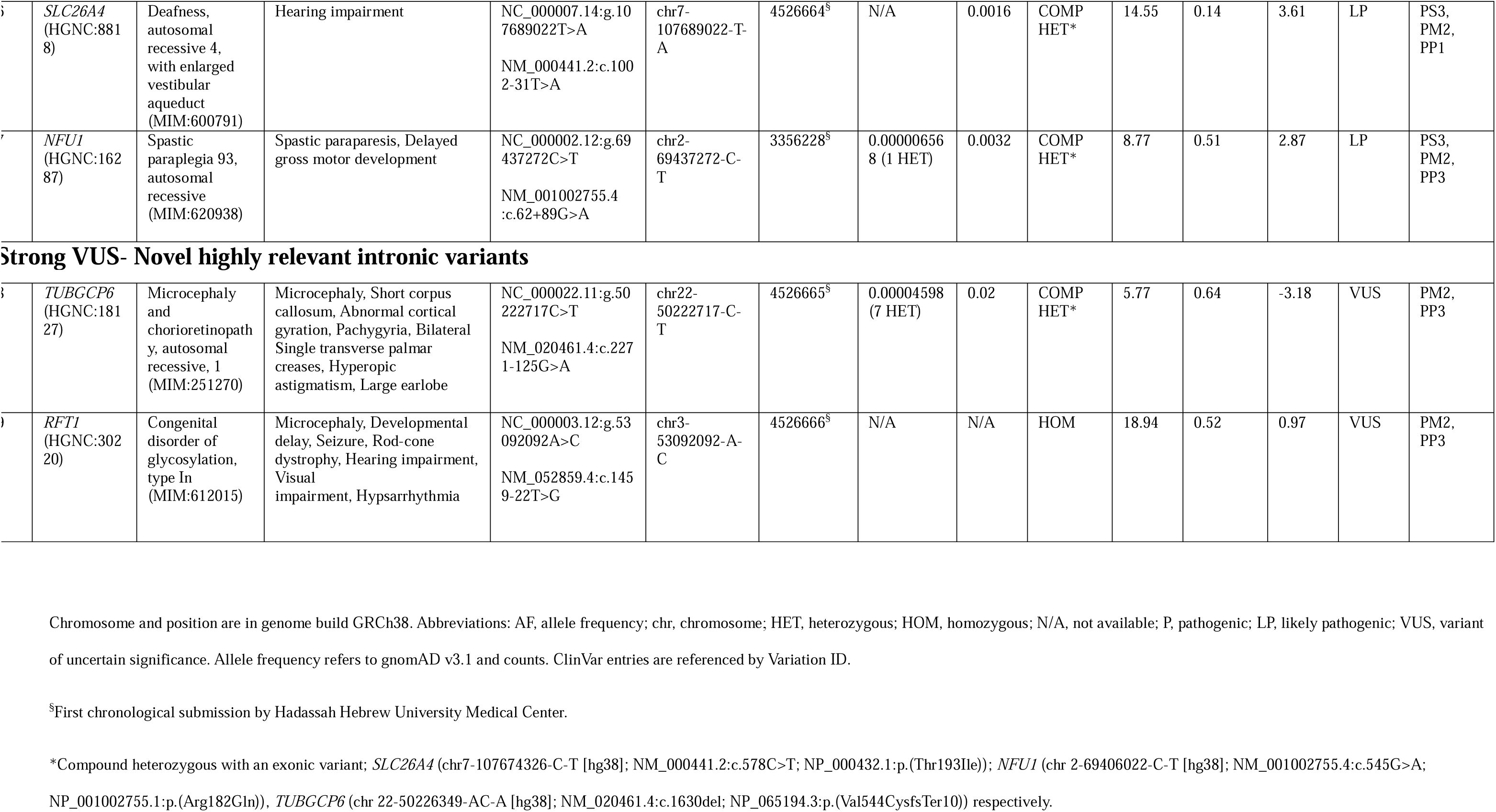
Reported variants identified using SMART Filtering.

| No. | Gene (HGNC ID) | Condition (OMIM) | Phenotype (HPO) | Genomic and Transcript HGVS (canonical) | Genomic position | ClinVar ID | Population AF (counts) | dbSNP | Zygosity | CADD | SPLICE-AI | GERP RS | ACMG final classification | ACMG supporting criteria |
|---|---|---|---|---|---|---|---|---|---|---|---|---|---|---|
| **Positive- Previously identified pathogenic variants** |  |  |  |  |  |  |  |  |  |  |  |  |  |  |
| 1 | *RNU4-2* (HGNC:10193) | ReNU syndrome (MIM:620851) | Intellectual disability, Seizure, Microcephaly, Talipes equinovarus, High palate | NC_000012.12:g.120291839_120291840insA<br>NR_003137.3:n.64_65insT | chr12-120291839-T-TA | 3068742 | N/A | N/A | HET | 20.8 | 0.05 | N/A | P | PS4, PS3, PM2 |
| 2 | *RNU2-2* (HGNC:10152) | N/A | Epileptic encephalopathy, Global developmental delay, Hypotonia | NC_000011.10:g.62841806C>T<br>NR_199791.1:n.4G>A | chr11-62841806-C-T | 3898494 | N/A | N/A | HET | 14.44 | 0.05 | 2.58 | P | PS4, PM2 |
| 3 | *ABCD4* (HGNC:68) | Methylmalonic aciduria and homocystinuria, cblJ type AR (MIM:614857) | Hypopigmentation of the skin, Hyperpigmentation of the skin, White hair, Depigmentation of skin, Hyperhomocysteinemia, Decreased circulating vitamin B12 concentration, Megaloblastic anemia, Erythroid dysplasia, Dyserythropoiesis, Pancytopenia, Methylmalonic acidemia | NC_000014.9:g.74286541C>T<br>NM_005050.4:c.1753-12G>A | chr14-74286541-C-T | 1701235 | N/A | N/A | HOM | 22.5 | 0.46 | -2.65 | LP | PM2, PM3 , PP1, PP3, PP4 |
| **Positive- Novel functionally validated intronic variants** |  |  |  |  |  |  |  |  |  |  |  |  |  |  |
| 4 | *PAX3* (HGNC:8617) | Waardenburg syndrome, type 1 and 3 (MIM:193500, MIM:148820) | Hypopigmentation of hair, Sensorineural hearing impairment, Clinical Waardenburg syndrome | NC_000002.12:g.222242699T>C<br>NM_181458.4:c.587-10416A>G | chr2-222242699-T-C | 4526662 | N/A | N/A | HET | 20.5 | 0.59 | -4.53 | LP | PS3, PM2, PP3, PP1 |
| 5 | *SERAC1* (HGNC:21061) | 3-methylglutaconic aciduria with deafness, encephalopathy, and Leigh-like syndrome (MIM:614739) | Developmental regression, Developmental delay, Microcephaly, Seizures, Hearing loss, Axial hypotonia, Appendicular spasticity, Decreased liver function, 3 Methyl-glutaric aciduria, 3-Methylglutaconic aciduria, Abnormal basal ganglia MRI signal intensity | NC_000006.12:g.158117431_158117432del<br>NM_032861.4:c.1403+297_1403+298del | chr6-158117428-CTT-C | 4526663<sup>§</sup> | N/A | N/A | HOM | 25.2 | 0.81 | N/A | LP | PS3, PM2, PP3 |
| 6 | *SLC26A4* (HGNC:8818) | Deafness, autosomal recessive 4, with enlarged vestibular aqueduct (MIM:600791) | Hearing impairment | NC_000007.14:g.107689022T>A<br>NM_000441.2:c.1002-31T>A | chr7-107689022-T-A | 4526664<sup>§</sup> | N/A | 0.0016 | COMP HET* | 14.55 | 0.14 | 3.61 | LP | PS3, PM2, PP1 |
| 7 | *NFU1* (HGNC:16287) | Spastic paraplegia 93, autosomal recessive (MIM:620938) | Spastic paraparesis, Delayed gross motor development | NC_000002.12:g.69437272C>T<br>NM_001002755.4:c.62+89G>A | chr2-69437272-C-T | 3356228<sup>§</sup> | 0.000006568 (1 HET) | 0.0032 | COMP HET* | 8.77 | 0.51 | 2.87 | LP | PS3, PM2, PP3 |
| **Strong VUS- Novel highly relevant intronic variants** |  |  |  |  |  |  |  |  |  |  |  |  |  |  |
| 8 | *TUBGCP6* (HGNC:18127) | Microcephaly and chorioretinopathy, autosomal recessive, 1 (MIM:251270) | Microcephaly, Short corpus callosum, Abnormal cortical gyration, Pachygyria, Bilateral Single transverse palmar creases, Hyperopic astigmatism, Large earlobe | NC_000022.11:g.50222717C>T<br>NM_020461.4:c.2271-125G>A | chr22-50222717-C-T | 4526665<sup>§</sup> | 0.00004598 (7 HET) | 0.02 | COMP HET* | 5.77 | 0.64 | -3.18 | VUS | PM2, PP3 |
| 9 | *RFT1* (HGNC:30220) | Congenital disorder of glycosylation, type In (MIM:612015) | Microcephaly, Developmental delay, Seizure, Rod-cone dystrophy, Hearing impairment, Visual impairment, Hypsarrhythmia | NC_000003.12:g.53092092A>C<br>NM_052859.4:c.1459-22T>G | chr3-53092092-A-C | 4526666<sup>§</sup> | N/A | N/A | HOM | 18.94 | 0.52 | 0.97 | VUS | PM2, PP3 |

Chromosome and position are in genome build GRCh38. Abbreviations: AF, allele frequency; chr, chromosome; HET, heterozygous; HOM, homozygous; N/A, not available; P, pathogenic; LP, likely pathogenic; VUS, variant of uncertain significance. Allele frequency refers to gnomAD v3.1 and counts. ClinVar entries are referenced by Variation ID.
<sup>§</sup>First chronological submission by Hadassah Hebrew University Medical Center.
\*Compound heterozygous with an exonic variant; *SLC26A4* (chr7-107674326-C-T [hg38]; NM_000441.2:c.578C>T; NP_000432.1:p.(Thr193Ile)); *NFU1* (chr 2-69406022-C-T [hg38]; NM_001002755.4:c.545G>A; NP_001002755.1:p.(Arg182Gln)), *TUBGCP6* (chr 22-50226349-AC-A [hg38]; NM_020461.4:c.1630del; NP_065194.3:p.(Val544CysfsTer10)) respectively.

Overall, SMART filtering increased the diagnostic yield by 4.6% (7/153), identified strong VUS in an additional 1.3% (2/153) of cases, and lower-certainty VUS in 5.2% of cases (8/153) as potential diagnoses.

### Candidate burden and ranking performance

We compared the performance of the standard GS pipeline (coding regions, ‘exons-only’ BED file; spanning ±50 bp around exonic boundaries) with the SMART pipeline (‘all-genes’ BED file with SMART pre-annotation filtering), across the reported and VUS variants within the candidate list. Both workflows used the same preset automatic filtering and manual review process, prior to the variant ranking comparison. In the standard pipeline, total variant count per case ranged from 39,620 to 45,735 (average: 41,281), with 1–85 variants remaining after applying preset-automatic filters. In SMART, the total and remaining variants after preset-automatic filters were higher (range: 36,038–87,193; average: 61,833 and 19-2,404 remaining variants per case), reflecting a genome-wide BED file.

Across all 153 cases analyzed using SMART, final post-filtering candidate sets contained 0–2,404 variants per case. After grouping variants by gene, the manual review burden was reduced to a median of 70 distinct candidate genes per case (IQR, 24–156.75; range, 0–1,057).

Using SMART, the nine diagnostic variants generally ranked within the first or second decile of the candidate list; in four of nine probands the diagnostic variant ranked first. By comparison, the standard pipeline missed the diagnostic variant in 4 of 9 cases, because the causative variants (*RNU4-2, PAX3, SERAC1*, *TUBGCP6*) were deep-intronic, residing outside the exonic BED file. The median normalized rank (rank ÷ total candidates after filtering) was 0.065, indicating typical placement within the top 6.5% of candidates, compared with 0.260 in the standard pipeline, among the 5 recoverable cases, an approximately four-fold improvement. Ranking data are provided in Table S3.

### Positive cases

#### Previously identified pathogenic variants

##### RNU4-2 n.64_65insT and *RNU2-2* n.4G>A

Individual 1 had bilateral clubfoot and microcephaly on a prenatal ultrasonography (Table 1). He was born at term and never achieved motor milestones. Later, he became wheelchair-dependent, non-verbal, and had epilepsy. Additional phenotypic features included bilateral clubfoot and a high palate. SMART filtering identified a heterozygous *RNU4-2* variant (chr12-120291839-T-TA [hg38]; NR_003137.3:n.64_65insT), a recurrent single-base insertion associated with a severe neurodevelopmental syndrome (MIM:620851). This variant disrupts spliceosome function by altering 5′ splice-site recognition and was recently identified in 89 individuals with unexplained neurodevelopmental delay (NDD), further validated by RNA-seq (Chen et al. 2024).

Individual 2 was evaluated for global developmental delay, refractory seizures, and status epilepticus, with hypotonia and absent speech. Prior ES (with multiple re-analyses) and clinical GS were negative. A heterozygous intronic *RNU2-2* variant (chr 11-62841806-C-T [hg38]; NR_199791.1:n.4G>A) with a CADD score of 14.44 was detected through SMART (Table 1). This variant was recently linked to a neurodevelopmental disorder (Greene et al. 2024). Segregation and further workup were unavailable.

Both individuals harbored previously reported pathogenic variants, serving as positive controls for the SMART approach and demonstrating its ability to identify clinically relevant variants and its utility for discovering novel findings.

##### *ABCD4* c.1753-12G>A

A female presented with patchy skin and hair pigmentation. Prior to genetic testing initiation, she was admitted to the pediatric intensive care unit (PICU) with pancytopenia and severe megaloblastic anemia, requiring blood transfusions. Biochemical evaluation revealed low vitamin B12, with elevated homocysteine, methylmalonic acid (MMA), and methionine. A similarly affected sibling was noted. A pedigree is provided in Fig S1.

SMART filtering identified a homozygous *ABCD4* intronic variant (chr14-74286541-C-T [hg38]; NM_005050.4:c.1753-12G>A), which was also found in a homozygous state in her affected sibling. This rare variant impacts a conserved nucleotide (CADD 22.5) and is predicted to alter splicing (SpliceAI 0.46) (Individual 3; Table 1). It has previously been reported in a compound heterozygous state in an individual with *ABCD4*-related biochemical profile (methylmalonic acidemia and homocystinuria). *ABCD4*-related disease may include feeding difficulties, failure to thrive, hypotonia, seizures, developmental delay, and hematological abnormalities (Pillai et al. 2021). Following the genetic diagnosis, clinical management was adjusted to include escalation of hydroxocobalamin therapy aimed at improving metabolic control.

#### Novel functionally validated intronic variants

##### *PAX3* c.587-10416A>G

A male presented with features and family history consistent with an autosomal dominant (AD) Waardenburg syndrome. Previous studies indicated linkage to the *PAX3* gene (MIM:193500, MIM:148820), yet sequencing of the gene and Multiplex Ligation-dependent Probe Amplification (MLPA) were negative. SMART filtering revealed a deep-intronic variant in intron 4 of *PAX3* (chr 2-222242699-T-C [hg38]; NM_181458.4:c.587-10416A>G) (Individual 4; Table 1 and Fig 2A). Conservation and prediction scores (GERP RS 4.53, CADD 20.50, SpliceAI 0.59) supported an effect on splicing.

**Figure 2.**
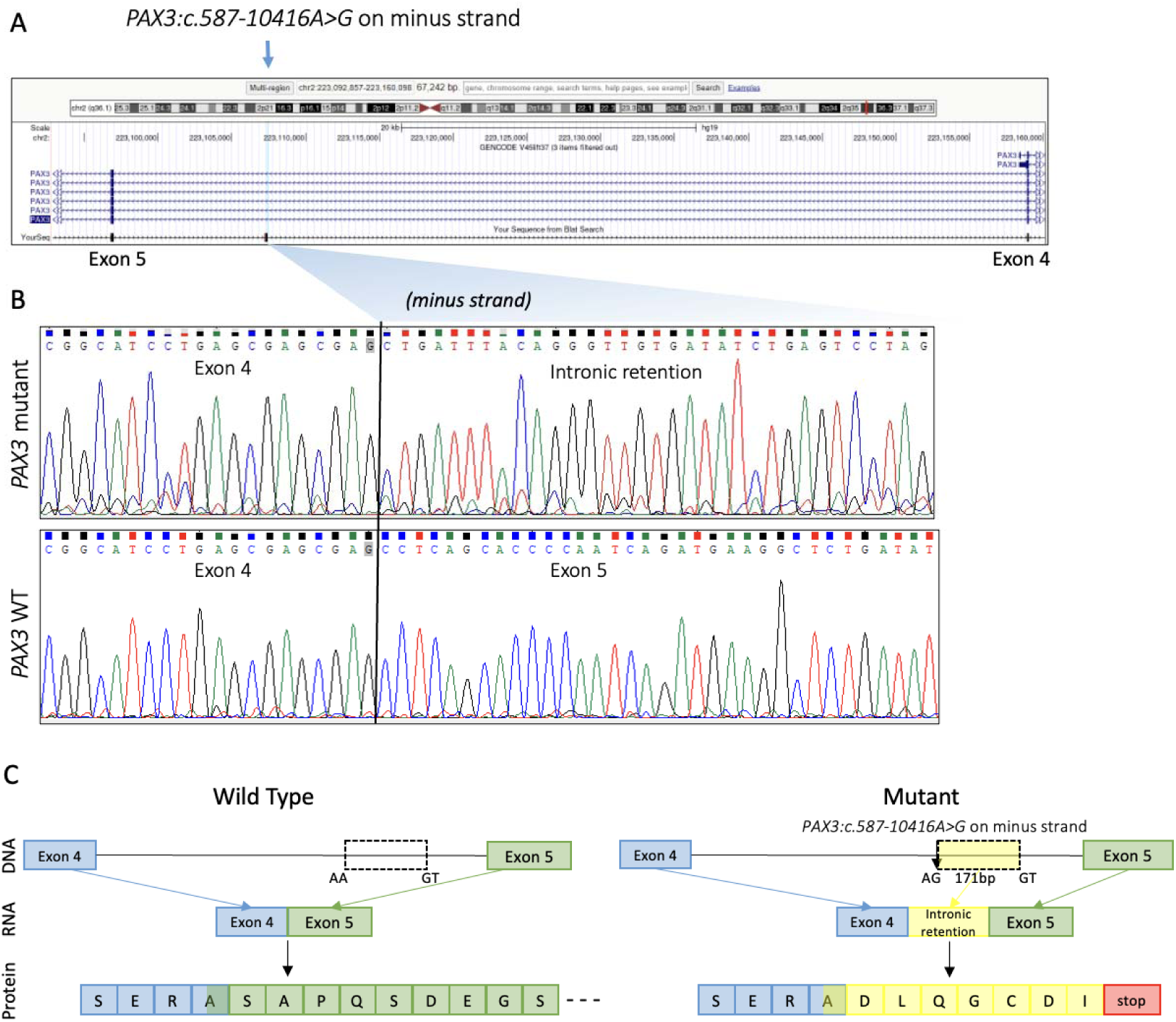
*PAX3* c.587-10416A>G variant laboratory validation process and results. (A) A deep-intronic variant was identified by SMART filtering GS analysis in intron 4 of the *PAX3* gene (*PAX3* c.587-10416A>G) in a patient with suspected Waardenburg syndrome. (B) RT-PCR followed by Sanger sequencing indicated that the variant generates a new splice site deep within intron 4, leading to the inclusion of a 171bp pseudo-exon in the mutant transcript. (C) In silico analysis further revealed the presence of a premature termination codon eight residues into the pseudo-exon sequence.

On the RNA level, RT-PCR from the patient’s fibroblasts showed reduced *PAX3* expression (Fig S2). CHX treatment revealed an elongated transcript in the affected sample; Sanger sequencing confirmed inclusion of a 171-bp pseudo-exon (Fig 2B, Fig S3), predicted to introduce a premature termination codon (Fig 2C) and reduce PAX3 expression through NMD.

The proposed pathogenic mechanism involves reduced *PAX3* expression, leading to a loss-of-function effect at the protein level. Considering the specific familial phenotype, supporting segregation, and the positive functional analysis, the final variant classification was LP (Table 1). The molecular diagnosis enabled approval for preimplantation genetic testing.

##### *SERAC1* 1403+297_1403+298del

The proband presented with developmental regression, seizures, severe feeding difficulties and failure to thrive (FTT), mild-moderate sensorineural hearing impairment, abnormal liver enzymes and axial hypotonia with peripheral hypertonia. *SERAC1*-associated disease (MIM:614739) was suspected given 3-methylglutaric and 3-methylglutaconic aciduria and characteristic brain magnetic resonance imaging (MRI) findings. Biallelic *SERAC1* variants are a known cause of 3-methylglutaconic aciduria with deafness, encephalopathy, and Leigh-like syndrome (MIM:614725) (Maas et al. 2017). Following negative ES and GS, SMART filtering identified a homozygous intronic indel in *SERAC1* (chr6-158117428-CTT-C [hg38]; NM_032861.4:c.1403+297_1403+298del) with a SpliceAI score of 0.8, highly suggestive of a splice altering effect (Individual 5; Table 1). The variant created a donor site motif (Fig 3A and Fig S4). RNA-Seq identified five aberrant transcripts in the variant region, as the predominant aberrant transcript, which accounted for 69% of reads, extended exon 13 by 387 nucleotides. Other aberrant transcripts included exon skipping and pseudo-exon activation (Petersen et al. 2022), while the normal transcripts observed in control samples were absent (Fig 3B-C and Fig S5, Table S4). These transcripts contained premature termination codons, collectively indicating a loss-of-function mechanism (Fig S6). Taken together, the variant was classified as LP.

**Figure 3.**
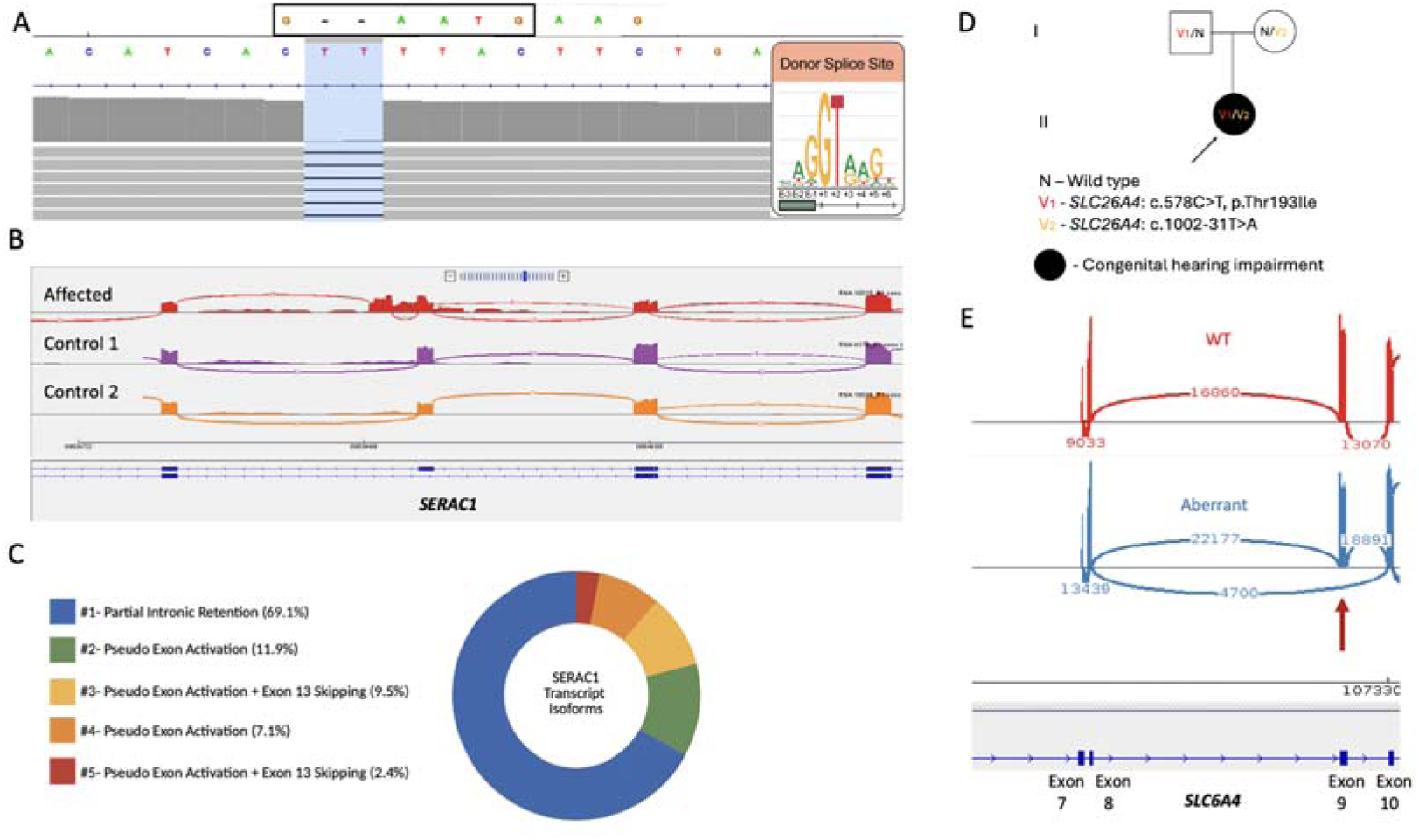
Functional Impact of the *SERAC1* c.1403+297_1403+298del and *SLC26A4* c.1002-31T>A variants. (A) Schematic representation of the *SERAC1* gene, highlighting the location of the 1403+297_1403+298del variant within the intronic region. Additionally, the variant created a novel sequence consistent with a donor splice-site motif. (B) Sashimi plot of RNA-seq analysis showing altered transcripts in affected individual compared to controls. (C) Pie chart illustration of the quantification of *SERAC1* variant transcript isoforms from RNA sequencing, demonstrating the variable splicing patterns caused by the variant. (D) Pedigree of the family with *SLC26A4* variants. Filled symbols denote affected individuals. Genotypes are N (wild-type allele), V1 (c.578C>T, p.Thr193Ile), and V2 (c.1002-31T>A); the proband harboring V1/V2 is indicated. (E) Sashimi plot presenting exon junctions for *SLC26A4* exon7-exon10 cDNA PCR product for a negative control (WT, red) and the heterozygous c.1002-31T>A variant in the patient (Aberrant, blue). Red arrow indicates exon 9 skipping. Numbers on splice junctions’ arches indicate the number of reads supporting the junction.

##### *SLC26A4* c.1002-31T>A

A female presented with congenital hearing impairment. She carried a single, known paternally-inherited, pathogenic variant in *SLC26A4* (chr7-107674326-C-T [hg38]; NM_000441.2:c.578C>T; NP_000432.1:p.(Thr193Ile)) (Campbell et al. 2001; Pryor et al. 2005), which was identified through ES-based gene panel. *SLC26A4* is associated with an autosomal recessive (AR) deafness (MIM:600791). SMART filtering identified a maternally-inherited intronic variant (chr7-107689022-T-A [hg38]; NM_000441.2:c.1002-31T>A) with a relatively high CADD score of 14.55, (Individual 6, Table 1, Fig 3D). This combination suggested a compound heterozygous diagnosis of congenital hearing impairment, although the intronic variant had not previously been reported as disease causing. RNA studies for the maternally-inherited variant (c.1002-31T>A) demonstrated aberrant splicing with exon 9 skipping in approximately 17% of transcripts, resulting in the deletion of 148 nucleotides and a frameshift, predicted to lead to loss-of-function (Fig 3E). The aberrant transcript may have been underestimated if it triggered NMD. The same inherited variant was later identified in another patient with bilateral enlarged vestibular aqueducts and sensorineural hearing loss. In this individual, the variant was found *in trans* with a previously established pathogenic *SLC26A4* variant (chr7-107690170-CT-C [hg38]; NM_000441.2:c.1198del; NP_000432.1:p.(Cys400ValfsTer32)), providing further evidence for its clinical significance and supporting the classification as likely pathogenic.

##### *NFU1* c.62+89G>A

The proband was a male who presented with delayed gross motor development and spastic paraparesis (Individual 7; Table 1), while intellectual development was reportedly intact. SMART filtering joint analysis was conducted using the GS of the proband and available ES of both his parents (Fig 4A). The analysis identified 2 variants in *NFU1*. First, a known paternally-inherited, pathogenic variant (chr2-69406022-C-T [hg38]; NM_001002755.4:c.545G>A; NP_001002755.1:p.(Arg182Gln)), which was reported in both homozygous and compound heterozygous states in individuals with NFU1-related disorders (Cameron et al. 2011; de Souza et al. 2018). RT-PCR studies published in the medical literature have demonstrated that this variant affected mRNA splicing (Cameron et al. 2011). The second variant is a rare, maternally-inherited intronic variant (chr2-69437272-C-T [hg38]; NM_001002755.4:c.62+89G>A), located in intron 1 of *NFU1* and previously reported as a VUS (Benkirane et al. 2021; Stark et al. 2025). Computational prediction tools suggested it had a deleterious effect on mRNA splicing (SpliceAI 0.51). RNA-seq revealed that the paternal variant (c.545G>A) resulted in two distinct transcripts: the first transcript demonstrated skipping of exon 6 and was detected in approximately 11% of sequencing reads spanning the exon junction (Fig S7). The second transcript exhibited a concurrent skipping of exons 6 and 7, observed in approximately 6% of the reads (Fig S8). These aberrant transcripts were absent in control samples, and both resulted in a frameshift and introduced a premature termination codon, which is predicted to lead to loss-of-function through either NMD or protein truncation. RNA-seq analysis of the region containing the maternal variant (c.62+89G>A) revealed partial inclusion of intron 1 in approximately 87% of the reads. This inclusion resulted in the addition of 90 nucleotides to exon 1, introducing a premature termination codon (Fig 4B). These aberrant reads were found to be *in cis* with the suspected variant and were also identified in control samples, but at a minimal frequency (Fig 4C, Table S5).

**Figure 4.**
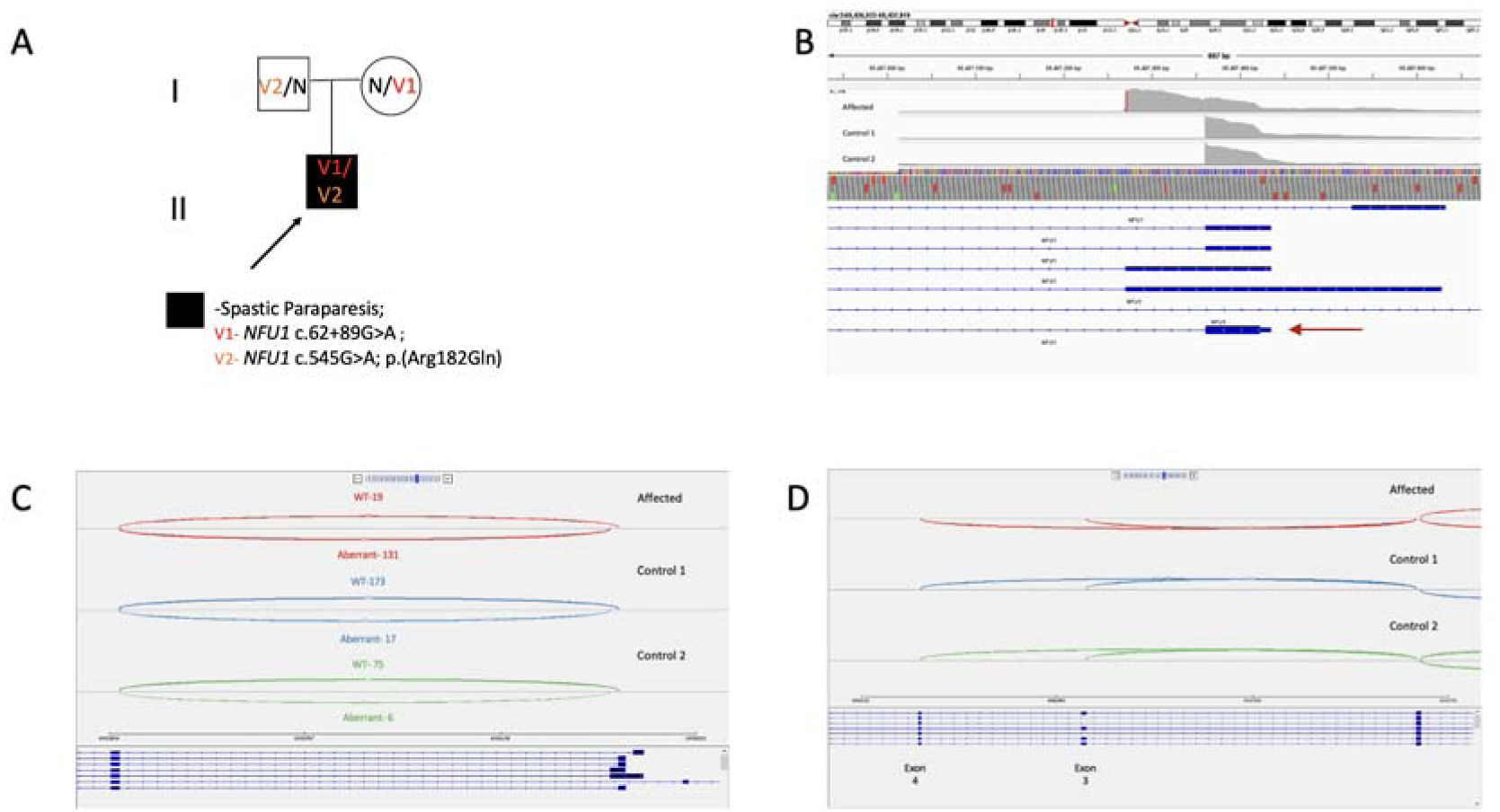
Identification of an intronic variant in *NFU1* and its effect on splicing. (A) Pedigree of the family with autosomal recessive spastic paraparesis due to compound heterozygous *NFU1* variants. Filled symbols indicate individuals with spastic paraparesis. Genotypes are annotated as N (wild-type *NFU1* allele), V1 (c.62+89G>A), and V2 (c.545G>A, p.Arg182Gln); the proband carrying V1/V2 is indicated by an arrow. (B) SMART filtering revealed an intronic variant: *NFU1* (chr2-69437272-C-T [hg38]; NM_001002755.4:c.62+89G>A), located in intron 1 of the gene. RNA-seq analysis demonstrated an abnormal transcript with partial inclusion of 90 nucleotides from intron 1 into exon 1 in the affected individual, compared with controls. The red arrow indicates the canonical *NFU1* transcript. (C) Sashimi plot illustrating the exon 1 junction of *NFU1* in the affected individual and 2 controls. In the affected sample, the c.62+89G>A intronic variant leads to partial inclusion of 90 nucleotides from intron 1 into exon 1, resulting in an aberrant transcript. Most reads spanning this region in the affected individual correspond to the aberrant transcript, whereas only minimal aberrant junction usage is detected in control samples. (D) Sashimi plot of *NFU1* RNA-seq data in the affected individual and 2 controls. In the affected sample, the plot demonstrates that c.62+89G>A intronic variant also leads to exon 3 skipping, which is consistent with alternative isoforms of this gene.

Additionally, transcripts with exon 3 skipping were increased relative to controls, consistent with the gene’s known alternative isoforms and the previously reported splicing effect of this variant. (Stark et al. 2025) (Fig 4D). Exon 3 skipping was also observed in control samples, but at lower ratios (Table S6).

Based on the phenotypic similarity and functional analysis, the variants were classified as likely pathogenic and were regarded as the patient’s diagnosis.‏

### Strong VUS cases

#### Novel highly relevant intronic variants

##### *TUBGCP6* c.2271-125G>A

An individual presented with microcephaly and abnormal prenatal ultrasonography findings, including short corpus callosum, abnormal cortical gyration and pachygyria with additional phenotypic features such as bilateral single transverse palmar creases, hyperopic astigmatism and large ears (Individual 8; Table 1). A similar phenotype was documented in a previous pregnancy. Previous CMA was unremarkable. Quad ES identified a maternally-inherited heterozygous *TUBGCP6* variant (chr22-50226349-AC-A [hg38]; NM_020461.4:c.1630del; NP_065194.3:p.(Val544CysfsTer10)) in both the proband and the affected fetus. The variant, classified as LP, was previously linked to AR developmental disorder characterized by microcephaly, delayed motor development and visual impairment, often accompanied by short stature (MIM:251270) (Martin et al. 2014). At the time of testing, it was present as a single heterozygous allele. A pedigree is provided in Fig S1.

SMART filtering identified an additional heterozygous variant in *TUBGCP6* (chr22-50222717-C-T [hg38]; NM_020461.4:c.2271-125G>A), with suggestive prediction scores (high SpliceAI 0.64, CADD 5.77), strongly suspected to be *in trans* with the maternal variant. The phasing of both variants could not be determined from short-read genome data. However, based on our internal database, these variants did not constitute a known haplotype, which supported a potential compound heterozygous inheritance pattern. Segregation and RNA studies were advised to assess the variant’s functional impact but had not yet been performed.

##### *RFT1* c.1459-22T>G

A female presented with severe global developmental delay, moderate hearing impairment (approx. -40dB), retinal degeneration resembling retinitis pigmentosa, and epilepsy characterized by hypsarrhythmia. Brain MRI revealed diffuse symmetric ventricular dilatation.

Prior testing including CMA, targeted analyses for *CDKL5* and Rett syndrome, research gene panels at other medical centers, and trio ES yielded negative results. Through SMART filtering on the GS data, a homozygous intronic variant in *RFT1* (chr3-53092092-A-C [hg38]; NM_052859.4:c.1459-22T>G) (Leroy 2006) was identified. Biallelic pathogenic variants in *RFT1* are associated with congenital disorder of glycosylation, type In, (MIM:612015). This variant had a high CADD score of 18.94 and was predicted to disrupt mRNA splicing with SpliceAI score of 0.52 (Individual 9; Table 1). Segregation analysis identified heterozygous carriers among unaffected family members, and RNA studies were recommended. A pedigree is provided in Fig S1.

## DISCUSSION

While no consensus method exists for GS analysis in previously negative cases, our findings indicate that adopting SMART filtering can improve diagnostic yield by broadening and systematically prioritizing the range of variants subjected to manual review. Among 153 patients with previously negative ES and clinical GS results, SMART filtering identified pathogenic or likely pathogenic variants in seven cases, increasing the diagnostic yield by 4.6% beyond the original GS analysis. It also identified strong VUS in two cases (1.3%) and lower certainty VUS in eight additional cases (5.2%).

In three cases, diagnoses relied on compound heterozygosity involving both coding and non-coding alleles, showing that SMART can effectively prioritize and identify both alleles even when they carry different functional predictions within the same genotype.

SMART filtering is particularly valuable in consanguineous populations, where homozygous variants are common, as demonstrated in the *SERAC1* and *ABCD4* cases. It may also prove useful when a single heterozygous pathogenic allele has already been identified, as in the *TUBGCP6*, *SLC26A4,* and *NFU1* cases. By prioritizing rare, non-coding, and regulatory variants, SMART makes it feasible to identify relevant variants among millions of non-coding changes. When applied to singleton GS, SMART may help compensate, at least in part, for the absence of parental data when trio sequencing, often limited by its high costs, is not available.

The ranking analysis provides a quantitative measure of SMART prioritization efficiency. Although it considers a broader search space (all genes rather than coding exons alone), its pre-annotation filters enrich for functionally relevant variants and improve the normalized rank (median 0.065 vs. 0.260 in the standard pipeline among recoverable cases). Most diagnostic variants fell within the first or second decile, reducing interpretation burden by placing causal variant within the top of the list. The four variants missed entirely by the standard pipeline due to their deep-intronic location (*RNU4-2, PAX3, SERAC1*, *TUBGCP6*), emphasize the limitations of exon-only restricted analysis and the value of a genome-wide approach such as SMART.

The SMART and standard pipelines differ in both search space and pre-annotation filtering, so their individual contributions to the ranking result cannot be separated in this analysis. A broader search space alone would be expected to worsen ranking because it enlarges the candidate pool; thus, the improved ranking with SMART reflects the pre-annotation filtering step. The benefit of the broader search space itself appears in sensitivity: four of the nine diagnostic variants were deep intronic and would have been missed entirely by the exon-only standard pipeline.

In our sample, 8 cases had a variant with lower certainty classified as VUS, for which the available evidence was insufficient to warrant further investigation at this time point. This highlights the need for larger genomic reference datasets and improved prediction tools, especially for non-coding regions. Additionally, our rarity filters relied partly on a local cohort, which may have introduced a population bias; we mitigated this by requiring rarity in both the local cohort and gnomAD. Therefore, further integration of data from healthy populations may help reduce the proportion of variants classified as VUS, a persistent challenge in genomic medicine.

This study has several limitations, most notably the relatively small sample size. Expanding the application of this approach to larger cohorts may enable a more accurate assessment of its diagnostic yield and broader clinical utility. Additionally, functional validation is often required to confirm the pathogenicity of identified variants in non-coding regions. RNA-seq remains the standard method of validation; however, this approach poses challenges when fresh tissues are unavailable or when the implicated gene is not expressed in accessible tissues. Moreover, transcript-level analyses can be further complicated by NMD, which may reduce or eliminate the mutant transcript, making detection and interpretation more difficult. Future studies may consider collecting patient samples prospectively to allow RNA-based analyses when needed, which could become a preferred strategy for variant interpretation (Lunke et al. 2023).

It is also important to note that part of the observed yield likely reflects advances in gene discovery and interpretation since the original analyses, including recognition of recently described *RNU* variants. Our results therefore reflect the combined effect of the SMART framework and the continually expanding genomic knowledge base.

While SMART filtering effectively prioritizes rare variants, it may fail to capture pathogenic variants with relatively high population frequencies or other variants with complex contributions to disease. Notably, this study did not include copy number variation (CNV) or structural variant (SV) analyses, as these were addressed separately during the initial clinical assessment. Nevertheless, the current limitations in CNV and SV prediction highlight the need for improved computational tools to accurately detect and interpret such variants.

Finally, as the cohort was assembled over several years spanning a laboratory-wide transition between GRCh37 and GRCh38, a mixed-assembly design was employed; while all annotations were applied in an assembly-aware manner, this may have had a limited impact on variant detection in assembly-discordant genomic regions.

In conclusion, SMART filtering provides a systematic and efficient framework for enhancing the diagnostic yield of rare disorders. Broader implementation of this approach, in combination with more comprehensive genomic databases and refined predictive algorithms, is expected to further improve diagnostic accuracy and patient outcomes. Future research should aim to establish standardized guidelines for the interpretation of non-coding variants to ensure consistency and reliability across studies and diagnostic laboratories.

## ACKNOWLEDGMENTS

The authors wish to thank the families for their participation in this study. They also wish to thank Arcensus Diagnostics for contributing with their expertise to defining the SMART filtering approach.

This work was performed as part of the requirements for Kamila Lee’s medical degree (MD) thesis on behalf of the Faculty of Medicine in the Hebrew University of Jerusalem.

## FUNDING STATEMENT

This research received no specific grant from any funding agency in the public, commercial, or not-for-profit sectors.

## CONFLICTS OF INTEREST

H.M.-S., Y.M., Y.U. are employees of Geneyx Genomics. Other authors declare no conflict of interest.

## AUTHOR CONTRIBUTION STATEMENT

H.M.-S., R.B., and K.L., designed the study, analyzed the samples and wrote the paper. T.H. supervised the study and the RNA validation studies. H.M.-S., Y.M., and Y.U developed the SMART filtering methods. S.Y-D. and I.G. conducted RNA studies and contributed to writing the paper. O.M., R.B., M.S., conducted RNA studies. N.S.L., M.C., L.G., O.F., T.K., T.D., D.R., R.R., B.P-S., H.S, and S.J. provided clinical data, and provided genetic counseling.

## DATA AVAILABILITY STATEMENT

All analyzed data contain patients’ personal data and are stored in accordance with institutional regulations. Pseudonymized data are available upon request. Classified variants were submitted to ClinVar (SUB15734851 and SUB15760295).

## ETHICAL APPROVAL

The study was conducted in accordance with IRB-approved protocol 0306-10-HMO.

## Notes

### Author Declarations

Ethics committee/IRB of Hadassah Medical Center gave ethical approval for this work 0306-10-HMO.

