## Supplementary file for "A Framework for Variant Filtering to Improve Diagnostic Outcomes in Genome Sequencing"

* These authors contributed equally

**Corresponding author:**

Hagar Mor-Shaked

+(972)-2-6777618 (fax)

**Human Genetics Journal**

**SUPPLEMENTARY MATERIALS AND METHODS**

**SMART filtering**

The SMART filtering application for previously unsolved cases was integrated into the Geneyx analysis platform. It includes a BED file ('all-genes') and a pre-annotation filtering step according to CADD score of >=5, SpliceAI >=0.049, and gnomAD V3.1 allele counts of 0 for homozygous and hemizygous variants and <=300 for heterozygous variants. Thresholds were determined experimentally, and according to the previous article published in 2023 [(26)](https://sciwheel.com/work/citation?ids=15704682&pre=&suf=&sa=0&dbf=0) addressing recommendations for intronic variant identification. The 'all-genes' BED file contains all genes in the human genome as reflected in ncbiRefSeq.txt.gz file, from txStart to txEnd + padding of 5,000bp on each side to include promoters and other regulatory regions. If padding of txStart leads to a negative number, the beginning of the gene is set to 0. When there are multiple transcripts per gene, the minimal txStart and maximal txEnd are taken, not necessarily from the same transcript. Genes on chromosomes 1-22, X, Y and M (mitochondria) were included. The file is generated separately for hg19 and hg38 and is available upon request.

To compare SMART filtering with a standard approach focused on coding regions, we generated an “all-exons” BED file. The files exome-default.hg19.bed and exome-default.hg38.bed were created using RefSeq gene annotations obtained from the UCSC Genome Browser (ncbiRefSeq.txt.gz, downloaded on June 12 and June 9, 2025, respectively). Only coding transcripts were retained. For each selected transcript, exon coordinates were extracted and extended by 50 base pairs at both the 5′ and 3′ ends to include flanking splice regions. The complete mitochondrial genome sequence was also included. All resulting intervals were subsequently combined, and overlapping regions were merged to generate a unified BED file.

**Variant annotation sources**

GRCh37:
ClinVar, 2025-06-08; dbSNP, b154; 1KGenome, 2013-05; ESP6500, 2; GERP, 2010; LRT, 4.4a; MutationTaster, 4.4a; SiftScore, 4.4a; UniProtAcc, 2025-06 ; MasterMind, 2025.04.19; SnpEff, v5.2b.gx3-2025-06; DbscSNV, 1.1; MitoMap, 2021-10; CADD, v1.7; SpliceAI, 1.3; GnomAD-exomes, 2.1.1; GnomAD-genomes, 2.1.1; MANE, 1.4; LitVar2, 2025-05; AlphaMissense, v2; Ensembl, t114; CIVIC, 2025-06-01; Omim, 2025-06; Entrez, 2025-06 ; DGV, v107_2020-02-25; DgvGold, v107_2016-05-15; GnomAD SV, v2.1; RmskPercent, 2022-10; CytogeneticSv, 2009-06; ClinGen, 2024-12; Camouflaged Genes, V1; Dark Genes, V1; Encode, 2025-06; SFARI, 2025-06;

GRCh38:
ClinVar, 2025-06-08; dbSNP, b154; 1KGenome, 2019-02; ESP6500, 2; GERP, 2010; LRT, 4.4a; MutationTaster, 4.4a; SiftScore, 4.4a; UniProtAcc, 2025-06; MasterMind, 2025.04.19; SnpEff, v5.2b.gx3-2025-06; DbscSNV, 1.1; MitoMap, 2021-10; CADD, v1.7; SpliceAI, 1.3; GnomAD-exomes, 2.1.1; GnomAD-genomes, 3.1.2; GnomADv4-exomes, 4.1.0; GnomADv4-genomes, 4.1.0; MANE, v1.4; LitVar2, 2025-05; AlphaMissense, v2; Ensembl, t114; CoLoRSdb, v1.2.0; Omim, 2025-06; Enterz, 2025-06; CIVIC, 2025-06-01; DGV, v107_2020-02-25; DgvGold, v107_2020-03-02; GnomAD SV, v2.1; RmskPercent, 2022-10; CytogeneticSv, 2022-10; ClinGen, 2024-12; Camouflaged Genes, V1; Dark Genes, V1; Encode, 2025-06; CoLoRSdb-Sv, v1.2.0; SFARI, 2025-06.

**RNA studies**

For *PAX3* studies, total RNA was isolated from cultured fibroblasts of an affected individual and control by TRIzol reagent extraction. cDNA was reverse transcribed from 1μg RNA using the qScript cDNA Synthesis Kit (Quantabio) which includes Oligo (dT) and random primers.

The region spanning exons 3 through 6 of *PAX3* was amplified by PCR reaction using PCRBIO HS Taq Mix Red (PCR BIOSYSTEMS) with forward primer: PAX3_EX3_RNA_F1: 5’-GGCATGTTCAGCTGGGAAAT-3’ and reverse primer: PAX3_EX6_RNA_R1: 5’-ATGTGGGCTGGTAAGAGGTC-3’. The resultant fragments were separated by 1.5% (w/v) agarose gel electrophoresis and their sequence determined by Sanger sequencing.

For *SLC26A4* studies, RNA was extracted from peripheral blood collected in Tempus™ tubes using the Tempus™ Spin RNA Isolation Kit (Life Technologies), following the manufacturer’s protocol. Reverse transcription was performed on 1µg RNA using Impromll reverse transcription system (Promega), with random hexamers as primers according to the manufacturer’s instructions. The cDNA served as template for splicing analysis. RT-PCR was performed with primers spanning SLC26A4 exons 8-10, from exon 7 to exon 11 (*-* SLC26A4-ex7-F: TGATCGGTTTAGACACAAAATCC and SLC26A4-ex11-R: CAAGACCGACTTCTGCAAGG). PCR products were analyzed by electrophoresis on 2% agarose gels.

**RNA-seq data processing and analysis**

RNA-seq data processing was performed using OmicsDiscoveries RNAvisionTM v.1.1.0 platform, which includes alignment, feature counting, and detection of aberrant expression, aberrant splicing, and mono-allelic expression as described below.

RNA-seq raw data were aligned using the nf-core rnaseq pipeline v3.19.0 (31) with the option --aligner = star_salmon. This included read trimming with Trim Galore v0.6.10 and read alignment with STAR 2.7.11b using the - -twopassMode = BASIC parameter. Quality metrics were collected through Qualimap v2.3, RSeQC v5.0.2 (31), and SAMtools v1.21, and visualized with MultiQC v1.29.0.  Further quality control included annotated individual and sex verification using DROP v1.5.0 (32). Tissue verification was performed by correlating the expression read counts across samples within and between tissues.

Detection of aberrant expression was based on DROP’s aberrant expression module using the GRCh38 primary assembly from GENCODE release 48 as a reference genome. Read pairs that fell completely within exonic regions were counted using the summarizeOverlaps function from the GenomicAlignments (33) R package. Reads that fully overlapped more than one feature were assigned to each feature. Genes with a 95th percentile FPKM < 0.5 were filtered out. Expression outliers were found using OUTRIDER v1.26.3 (34).

Detection of aberrant splicing events was also based on DROP, which uses the Intronic Jaccard Index based on FRASER v2.4.6 (35), an annotation-free aberrant splicing detection algorithm. Junctions with < 20 reads in all samples and for which the total number of reads at the donor and acceptor splice site was 0 in more than 75% of the samples were filtered out.

Mono-allelic expression was performed by counting the reads assigned to each allele on positions with heterozygous single-nucleotide variants using the ASEReadCounter function from GATK v4.4 (36). Positions with less than 10 reads assigned to either allele in total were filtered out.

To increase statistical power, 95 additional samples with matching tissue and sequencing protocols were incorporated, jointly processed, and used in the combined model fitting.

Aberrant events were visually inspected through the RNAvisionTM platform and the Integrative Genomics Viewer (IGV).

**Phenotyper for phenotype prioritization**

**Overview**

Phenotype-driven candidate gene prioritization was performed using Geneyx Phenotyper, a proprietary algorithm developed and maintained within the Geneyx cloud infrastructure (not publicly available). The algorithm accepts any phenotype descriptor; including free-text clinical terms, disease names, or structured Human Phenotype Ontology (HPO) terms, and produces a ranked list of candidate genes by integrating variant-level, gene-level, and phenotype-level evidence.

**1. Data Acquisition, Versioning, and Normalization**

Source ingestion: Phenotyper integrates approximately 50 public and commercial databases, downloaded on a fixed 2–3-month cycle. These include:

1. HGNC genes, 2025-06
2. NCBI Gene to PubMed
3. NCBI Gene information
4. NCBI Gene to RefSeq
5. Entrez Genes Homo sapiens, 2025-06
6. PubMed publications
7. OMIM, 2025-06
8. HPO terms, 2025-06
9. Gene-to-HPO mappings, 2025-06
10. GeneImprint, 2025-06
11. ClinVar, 2025-06
12. Ensembl Genes (hg19/hg38)
13. RefSeq Genes (hg19/hg38), 2025-06
14. Orphanet (diseases, genes, and symptoms), 2025-06
15. UniProt, 2025-06
16. ENCODE, 2025-06
17. eQTL GTEx analysis, 2025-06
18. Array Express (promoter data)
19. DECIPHER, 2025-06
20. MGI
21. GTEx expression
22. Disease-specific gene-to-phenotype resources (cancer, cardiac, eye, skin, and others)
23. CPIC
24. CPIC Drugs
25. CPIC Activities
26. Diplotype Frequency
27. CPIC Interpretation
28. LitVar2, 2025-05
29. Geneyx Analysis curated data

Normalization pipeline: Each database undergoes a standardized ETL (extract–transform–load) process comprising schema harmonization into a unified SQL Server relational model; deduplication and conflict-resolution heuristics; ontology alignment to HPO and DOID; normalization of gene identifiers to HGNC symbols; and ElasticSearch indexing using tokenized free-text, multi-field normalization, and cross-field synonym expansion.

Reproducibility controls: All ETL jobs are orchestrated through Apache Airflow (v2.1+) with the following reproducibility properties: version pinning, whereby every database instance is stored with a timestamped version tag; parse verification, including row counts, null-distribution checks, and schema conformity logs; and index validation via ElasticSearch shard health and term-frequency diagnostics. All metadata, including database versions, update dates, and schema changes are preserved to ensure full analytical reproducibility.

**2. Algorithmic Structure and Scoring**

Operational modes: Phenotyper operates in two modes: *clinical mode*, which uses only curated data sources and strong, well-validated gene–phenotype associations; and *research mode*, which additionally incorporates weaker or preliminary associations.

**2.1 Direct Evidence Scoring**

A direct phenotype–gene score is computed from four evidence streams:

- Term-frequency / inverse-document-frequency (TF–IDF) matches between patient HPO terms and indexed gene-associated documents
- Matches in structured fields, including gene–disease tables, curated phenotype associations, and structured ontology links
- Matches in unstructured biomedical text following NLP normalization (stemming, synonym expansion, and negation detection)
- Weighted boost factors applied to fields with high clinical inference value (e.g., HPO term labels, disease names, publication titles)

Each match contributes a weighted score according to:

**DirectScore(gene) = Σᵢ wᵢ × TF–IDF(termᵢ, gene)**

where *wᵢ* is a field-specific weighting factor reflecting the clinical relevance of the matched field.

**3. Composite Prioritization Score and Interpretability**

For each candidate gene, the final prioritization score is:

**FinalScore = DirectScore**

The composite score is an unscaled continuous metric; it does not follow a normalized 0–100 or probability scale, but accumulates additively as matching evidence is identified across databases and fields. Based on internal validation, a score of ≥20 empirically corresponds to robust genotype–phenotype correlation, typically reflecting concordant support from multiple high-confidence curated sources (e.g., OMIM disease entries, direct HPO mappings, and ClinVar pathogenic associations). Scores below this threshold may still be informative when the patient's HPO profile is rare or incompletely represented in structured databases; accordingly, genes demonstrating alignment with the majority of the patient's HPO terms were also retained as candidates regardless of absolute score.

In addition to the score, each gene receives: a full evidence trace documenting all matched fields, sources, and seed phenotypes; field-level contribution breakdowns to support interpretability; and score stability across runs, ensured by version-fixed database snapshots.

**4. System Architecture and Reproducibility**

- Execution environment**:** Scoring runs inside a controlled, containerized environment to guarantee runtime consistency across analyses.
- Quality assurance: For every new database version, regression tests compare scoring outputs against the prior version to detect score drift or systematic changes.
- Reproducibility guarantee: Every analysis is tagged with the Phenotyper version in use; each version tag links to the exact database versions and scoring parameters applied. Any analysis can be fully regenerated by reloading the corresponding database snapshot.

| Individual | Genomic build | Total SNVs (VCF) | After REF/NO_CALL removal | After “all-genes” BED filter | BED retention (%) | After pre-annotation SMART filters |
| --- | --- | --- | --- | --- | --- | --- |
| Individual 3 (*ABCD4*) | GRCh38 | 4,908,455 | 4,907,326 | 4,071,148 | 83.0% | 87,193 |
| Individual 5 (*SERAC1*) | GRCh37 | 4,889,649 | N/R | 3,433,826 | 70.2% | 55,549 |
| Individual 8 (*TUBGCP6*) | GRCh38 | 4,987,371 | 4,986,866 | 4,162,830 | 83.5% | 86,950 |
| Individual 9 (*RFT1*) | GRCh38 | 4,966,474 | 4,965,354 | 4,175,144 | 84.1% | 87,021 |

**Table S1. Effect of pre‑annotation SMART filtering on variant counts**

In four representative genomes, showing total SNVs, variants intersecting the all-genes BED file, BED retention (%), and the number of variants remaining after SMART preset automatic filters application.

| No. | Gene (HGNC ID) | Condition (OMIM) | Phenotype (HPO) | Genomic and Transcript HGVS (canonical) | Genomic position | ClinVar ID | Population AF (counts) | dbSNP | Zygosity | CADD | SPLICE-AI | GERP-RS | ACMG final classification | ACMG supporting criteria |
| --- | --- | --- | --- | --- | --- | --- | --- | --- | --- | --- | --- | --- | --- | --- |
| 10 | *ETV6*  (HGNC:3495) | Leukemia, acute myeloid (MIM:600618) | Acute Myeloid Leukemia, Cancer, Leukemia | NC_000012.12:g.11863819T>C  NM_001987.5:c.464-5605T>C | chr12-11863819-T-C | N/A | 0.000006569 (1 HET) | 0.0008 | HET | 9.63 | 0 | 1.04 | VUS | PM2, BP7 |
| 11 | *ITPR3*  (HGNC:6182) | Charcot-Marie-Tooth disease, demyelinating, type 1J AD (MIM:147267) | Demyelinating peripheral neuropathy, Polyneuropathy, Sensorimotor polyneuropathy | NC_000006.12:g.33670827_33670845del  NM_002224.4:c.2586+12_2586+30del | chr6-33670817-TGGCTGGGGGAGTGCCCAGG-T | N/A | 0.000006573 (1 HET) | N/A | HET | 16.34 | 0.57 | N/A | VUS | PM2, BP7 |
| 12 | *GDAP1*  (HGNC:15968) | Charcot-Marie-Tooth disease (MIM:606598) | Myopathy, Lipid storage myopathy, Neuropathy, Von Willebrand disease, Muscle weakness | NC_000008.11:g.74360125T>G  NM_018972.4:c.311-12T>G | chr8-74360125-T-G | N/A | N/A | N/A | HET | 14.52 | 0.1 | 5.15 | VUS | PM2, BP7 |
| 13 | *C9orf72*  (HGNC:28337) | Frontotemporal dementia and/or amyotrophic lateral sclerosis 1, AD (MIM:614260) | Mental deterioration, Cognitive decline, Cerebral atrophy, Frontotemporal dementia-suspect, Unsteady gait | NC_000009.12:g.27571159_27571160del  NM_018325.5:c.-45+2275_-45+2276del | chr9-27571154-CAG-C | N/A | 0.00002629 (4 HET) | 0.0032 | HET | 3.55 | 0 | N/A | VUS | PM2, BP7 |
| 14 | *CTNNA2*  (HGNC:2510) | Cortical dysplasia, complex, with other brain malformations 9 AR (MIM:114025) | Microcephaly, Seizure, Global developmental delay, Large ears | NC_000002.12:g.80351482A>G  NM_001282597.3:c.1057-41729A>G | chr2-80351482-A-G | N/A | 0.00001315 (2 HET) | 0.0016 | HOM | 18.08 | 0.02 | 4.5 | VUS | PM2, BP7 |
| 15 | *CIT*  (HGNC:1985) | Microcephaly 17, primary, autosomal recessive AR (MIM:605629) | Short stature, Hypertonia, Patent ductus arteriosus, Seizures, Microcephaly, Intellectual disability, Dysmorphism, Perinatal asphyxia, Hypoxic ischemic encephalopathy, Rt hemiparesis, Global developmental delay, Seizures, Sleeping disturbance, Patent ductus arteriosus, Asymmetric facial features, Pear shaped nose, Upslanted palpebral fissures, Hirsutism, Thin Corpus callosum, Abnormal brain imaging with thin CC, Arachnoid cyst, Leukomalacia | NC_000012.12:g.119841020_119841021del  NM_001206999.2:c.517-6793_517-6792del | chr12-119841019-TAA-T | N/A | N/A | N/A | HOM | 0.415 | 0.05 | N/A | VUS | PM2, BP7 |
| 16 | *AUTS2*  (HGNC:14262) | Intellectual developmental disorder, autosomal dominant 26 AD (MIM:607270) | Microcephaly, Brachydactyly, Global developmental delay, Unilateral cryptorchidism, Triangular  face, Hypertelorism, Anteverted nares, Long philtrum, Thin upper lip vermilion | NC_000007.14:g.70389791T>C  NM_015570.4:c.661-45961T>C | chr7-70389791-T-C | N/A | N/A | N/A | HET | 20.7 | 0 | 5.21 | VUS | PM2, BP7 |
| 17 | *BRCA1*  (HGNC:1100) | Breast-ovarian cancer, familial, 1 AD (MIM:113705) | Breast carcinoma | NC_000017.11:g.43076853G>A  NM_007294.4:c.4358-239C>T | chr17-43076853-G-A | N/A | 0.000006598 (1 HET) | 0.0032 | HET | 8.63 | 0 | 1.85 | VUS | PM2, BP7 |

**Table S2.** **Lower certainty candidate variants detected using SMART Filtering.**

This table lists lower-certainty VUS variants identified through SMART filtering, including the affected gene, associated OMIM condition, HPO phenotype, HGVS and genomic positions (GRCh38), dbSNP, zygosity, CADD and SpliceAI scores, and ACMG classifications. Abbreviations: chr, chromosome; HET, heterozygous; HOM, homozygous; N/A, not available; P, pathogenic; LP, likely pathogenic; VUS, variant of unknown significance. Allele frequency refers to gnomAD v3.1 and counts. ClinVar entries were not available.

| Reported Variants | | | Default - Exons only | | | | SMART- All genes | | | |
| --- | --- | --- | --- | --- | --- | --- | --- | --- | --- | --- |
| Individual | Gene | VCF variants before BED | Total variants after BED file | Variants after applying preset-automatic filtering | Rank | Noramalized rank | Total variants after BED file+pre-filters | Variants after applying preset-automatic filtering | Rank | Noramalized rank |
| 1 | *RNU4-2* | 4827418 | 40139 | 72 | Undetected | N/A | 58477 | 2404 | 304 | 0.126 |
| 2 | *RNU2-2* | 4924062 | 40639 | 22 | 4 | 0.182 | 36038 | 690 | 104 | 0.151 |
| 3 | *ABCD4* | 4908455 | 40808 | 4 | 1 | 0.250 | 87193 | 19 | 1 | 0.053 |
| 4 | *PAX3* | 4810436 | 41111 | 2 | Undetected | N/A | 54168 | 76 | 1 | 0.013 |
| 5 | *SERAC1* | 4889649 | 40658 | 61 | Undetected | N/A | 55549 | 1283 | 1 | 0.001 |
| 6 | *SLC26A4* | 4972013 | 41632 | 12 | 4 | 0.333 | 85624 | 89 | 10 | 0.112 |
| 6 | *SLC26A4* | 4972013 | 41632 | 12 | 3 | 0.250 | 85624 | 89 | 9 | 0.101 |
| 7 | *NFU1* | 4846579 | 40362 | 3 | 2 | 0.667 | 38553 | 54 | 4 | 0.074 |
| 7 | *NFU1* | 4846579 | 40362 | 3 | 1 | 0.333 | 38553 | 54 | 3 | 0.056 |
| 8 | *TUBGCP6* | 4987371 | 41514 | 4 | Undetected | N/A | 86950 | 89 | 2 | 0.022 |
| 8 | *TUBGCP6* | 4987371 | 41514 | 4 | 1 | 0.250 | 86950 | 89 | 1 | 0.011 |
| 9 | *RFT1* | 4966474 | 42143 | 48 | 13 | 0.271 | 87021 | 1008 | 373 | 0.370 |
| VUS Variants | | | Default - Exons only | | | | SMART- All genes | | | |
| Individual | Gene | VCF variants before BED | Total variants after BED file | Variants after applying preset-automatic filtering | Rank | Noramalized rank | Total variants after BED file+pre-filters | Variants after applying preset-automatic filtering | Rank | Noramalized rank |
| 10 | *ETV6* | 4800748 | 41189 | 1 | Undetected | N/A | 49895 | 23 | 11 | 0.478 |
| 11 | *ITPR3* | 4852358 | 41368 | 9 | Undetected | N/A | 53583 | 69 | 57 | 0.826 |
| 12 | *GDAP1* | 4884769 | 41924 | 28 | Undetected | N/A | 54786 | 374 | 34 | 0.091 |
| 13 | *C9orf72* | 4906319 | 41862 | 13 | Undetected | N/A | 57728 | 225 | 5 | 0.022 |
| 14 | *CTNNA2* | 4600407 | 40372 | 75 | Undetected | N/A | 76719 | 1744 | 39 | 0.022 |
| 15 | *CIT* | 4857747 | 40697 | 85 | Undetected | N/A | 39602 | 1665 | 99 | 0.059 |
| 16 | *AUTS2* | 4758668 | 39620 | 26 | Undetected | N/A | 47303 | 617 | 46 | 0.075 |
| 17 | *BRCA1* | 5410705 | 45735 | 6 | Undetected | N/A | 81973 | 30 | 5 | 0.167 |

**Table S3. Candidate burden and diagnostic variant ranking**

Comparison between standard “exons-only” vs. SMART pipelines, showing variant counts before/after BED file restriction and preset automatic filtering. The rank and normalized rank of reported and VUS variants are shown.

| **Transcript #** | **Genomic Coordinates (GRCh37)** | **Genomic Coordinates (GRCh38)** | **Effect** | **% of reads** |
| --- | --- | --- | --- | --- |
| 1 | Chr6:158538467 - 158538853 | Chr6:158117435 - 158117821 | Partial intronic retention causing extension of exon 13 | 69.0% |
| 2 | Chr6:158538467 - 158538607 | Chr6:158117435 -158117575 | Pseudo exon activation | 11.9% |
| 3 | Chr6:158538467 - 158538607 | Chr6:158117435 -158117575 | Pseudo exon activation and exon 13 skipping | 9.5% |
| 4 | Chr6:158538467 -158538604 | Chr6:158117435 -158117572 | Pseudo exon activation | 7.1% |
| 5 | Chr6:158538467 - 158538604 | Chr6:158117435 -158117572 | Pseudo exon activation and exon 13 skipping | 2.4% |

**Table S4: Transcript isoform quantification and splicing effects of *SERAC1* 1403+297_1403+298delAA variant.**

This table presents the quantification of five distinct *SERAC1* variant transcript isoforms identified through RNA sequencing, illustrating the variable splicing patterns caused by the 1403+297_1403+298delAA variant.

| **Transcript Type** | **Genomic Coordinates (GRCh38)** | **Affected** | **Control 1** | **Control 2** | **Control 3** | **Control 4** |
| --- | --- | --- | --- | --- | --- | --- |
| Wild-type (exon 1 WT read) | Chr2:69432005-69437360 | 19 | 173 | 75 | 271 | 174 |
| Aberrant (exon 1 extension) | Chr2:67432005-69437270 | 131 | 17 | 6 | 5 | 4 |
| % extension / total | ---- | 87% | 8.9% | 7.4% | 1.8% | 2.2% |

**Table S5: Quantification of *NFU1* wild-type and aberrant splice junction reads- exon 1 extension.**
Number of RNA-seq reads spanning the canonical (WT) and aberrant splice junctions in the affected individual and four control samples. The c.62+89G>A variant in the affected individual leads to a clear predominance of aberrant junction reads, whereas controls show only minimal levels of this splicing event (up to 10%).

| **Transcript Type** | **Genomic Coordinates (GRCh38)** | **Affected** | **Control 1** | **Control 2** | **Control 3** | **Control 4** |
| --- | --- | --- | --- | --- | --- | --- |
| Wild-type (Exon 2-3 WT reads) | Chr2:69423718-69431901 | 229 | 427 | 209 | 306 | 162 |
| Aberrant (exon 3 skipping) | Chr2:69419605-69431901 | 161 | 89 | 31 | 38 | 50 |
| % skipping / total | ---- | 41% | 17% | 13% | 11% | 24% |

**Table S6: Quantification of *NFU1* wild-type and aberrant splice junction reads- exon 3 skipping.**
Number of RNA-seq reads spanning the canonical (WT) and aberrant splice junctions in the affected individual and four control samples. The affected individual shows a clear predominance of aberrant junction reads, whereas controls show relatively lower levels of this splicing event (~40% vs. up to 24%).

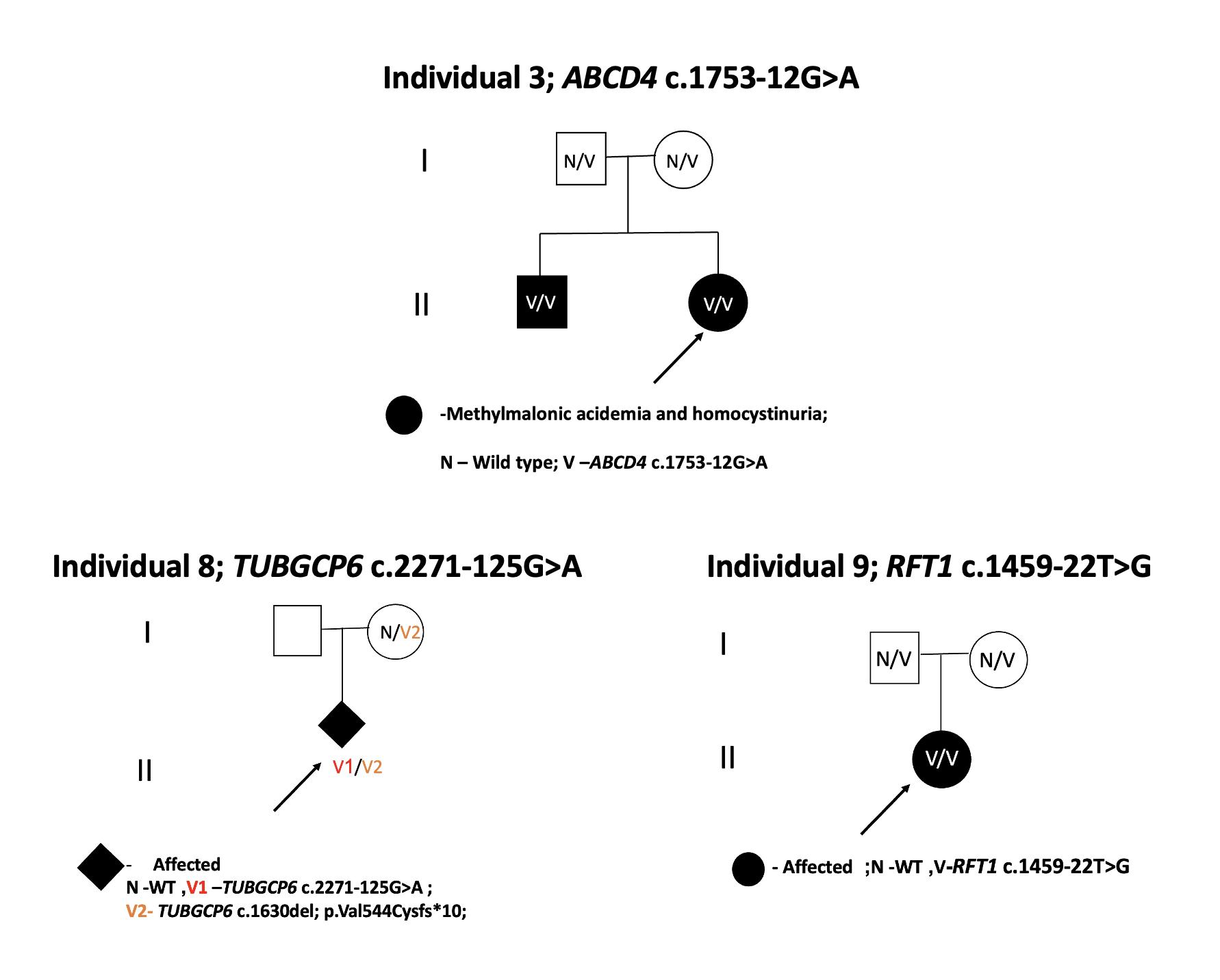

**Figure S1. Pedigrees of additional families included in the study.**

Pedigrees of participating families are included based on available consent. Filled symbols indicate affected individuals; open symbols indicate unaffected individuals. Genotypes are shown below each symbol (N, wild type allele; V or V1/V2, variant alleles as indicated). Clinical diagnoses are listed beneath the corresponding pedigrees.

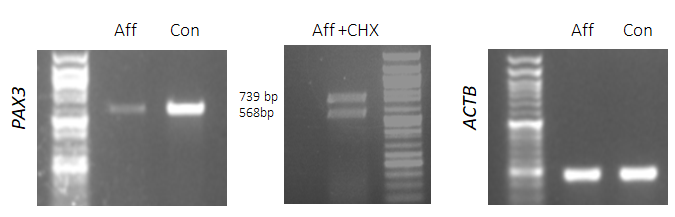

**Figure S2: *PAX3* RT-PCR results.**

Left panel: RT-PCR across intron 4 demonstrating reduced *PAX3* expression in fibroblasts derived from the affected individual compared to control. Middle panel: Cycloheximide (CHX) treatment prevented the degradation of aberrant transcripts, revealing a higher band representing a longer transcript in affected cells. Right panel: *ActinB* was used as a housekeeping gene. Aff - affected; Con - control.

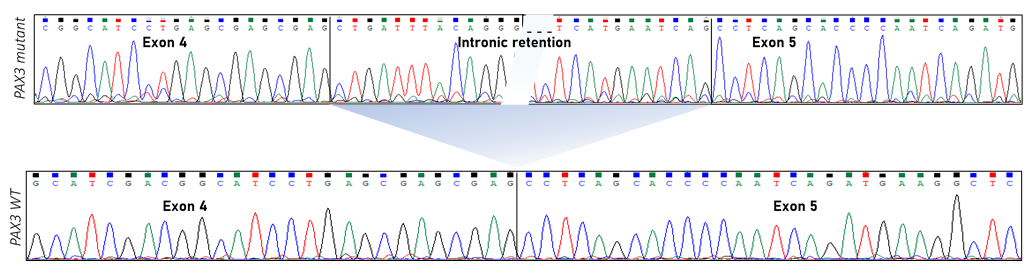

**Figure S3: Sanger sequencing confirmation.** Sanger sequencing results confirming the in-frame inclusion of a 171bp fragment from intron 4 in the *PAX3* mutant transcript.

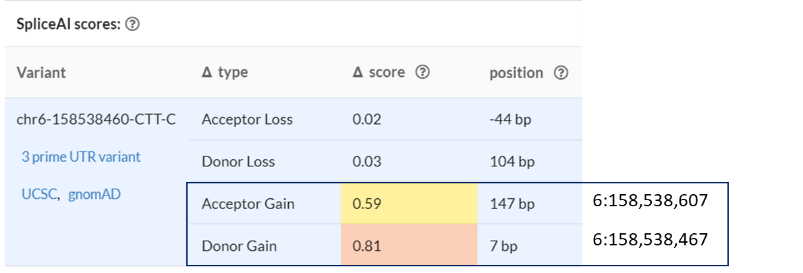

**Figure S4: SpliceAI Prediction tool results for *SERAC1* 1403+297_1403+298delAA** **variant.** The high scores predict donor gain at position 6:158538607 (hg19/GRCh37) and an acceptor gain at position 6:158538467 (hg19/ GRCh37), suggesting the potential creation of novel splice sites.

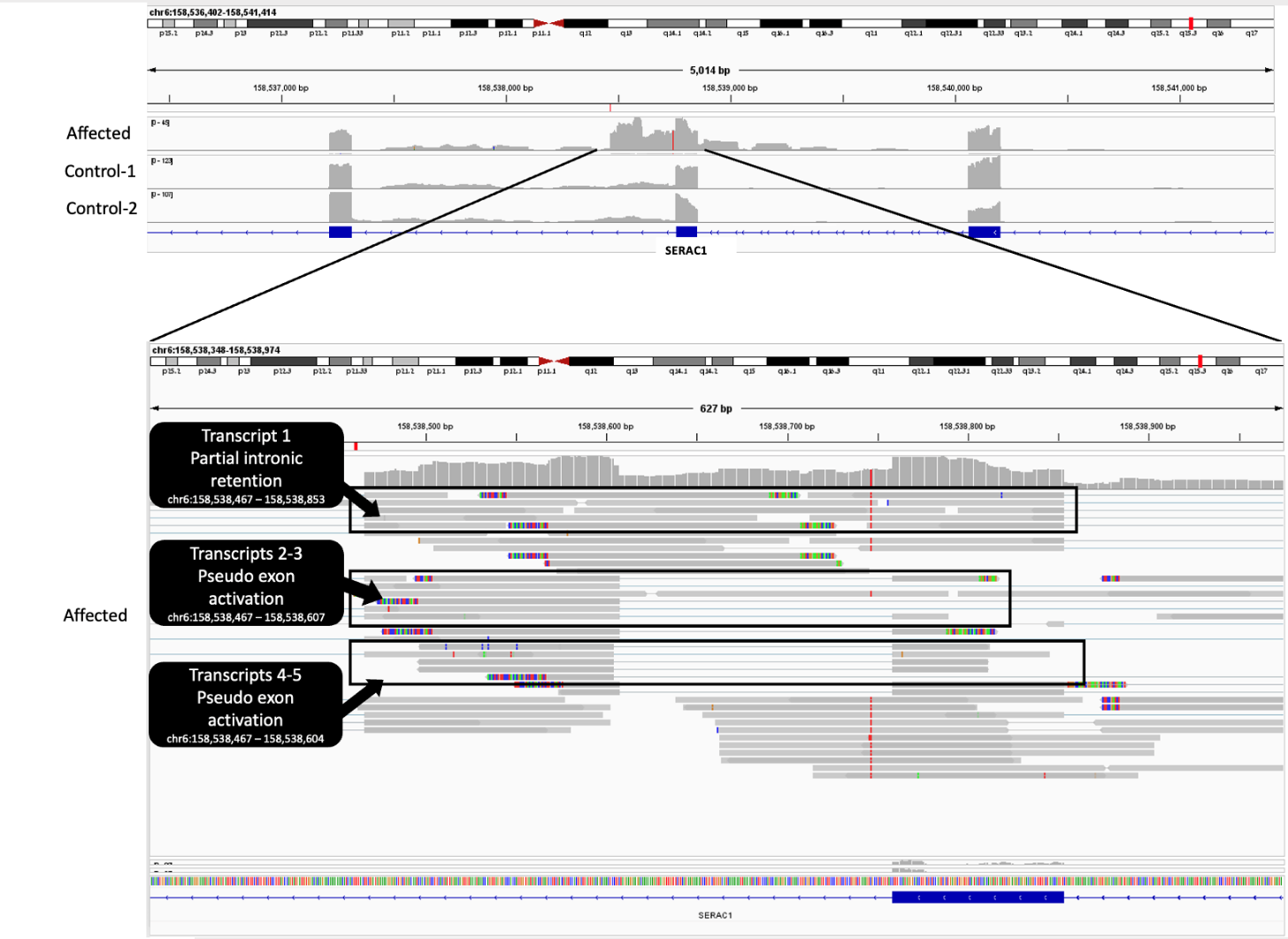

**Figure S5: Integrative Genomics Viewer (IGV) Visualization of the *SERAC1* Variant.** IGV visualization of RNA sequencing reads aligned to the *SERAC1* locus, highlighting the deletion at position 1403+297_1403+298 in red. The deletion is associated with altered splicing patterns, resulting in multiple transcript isoforms as compared to the controls. (Genomic positions in hg19/GRCh37)

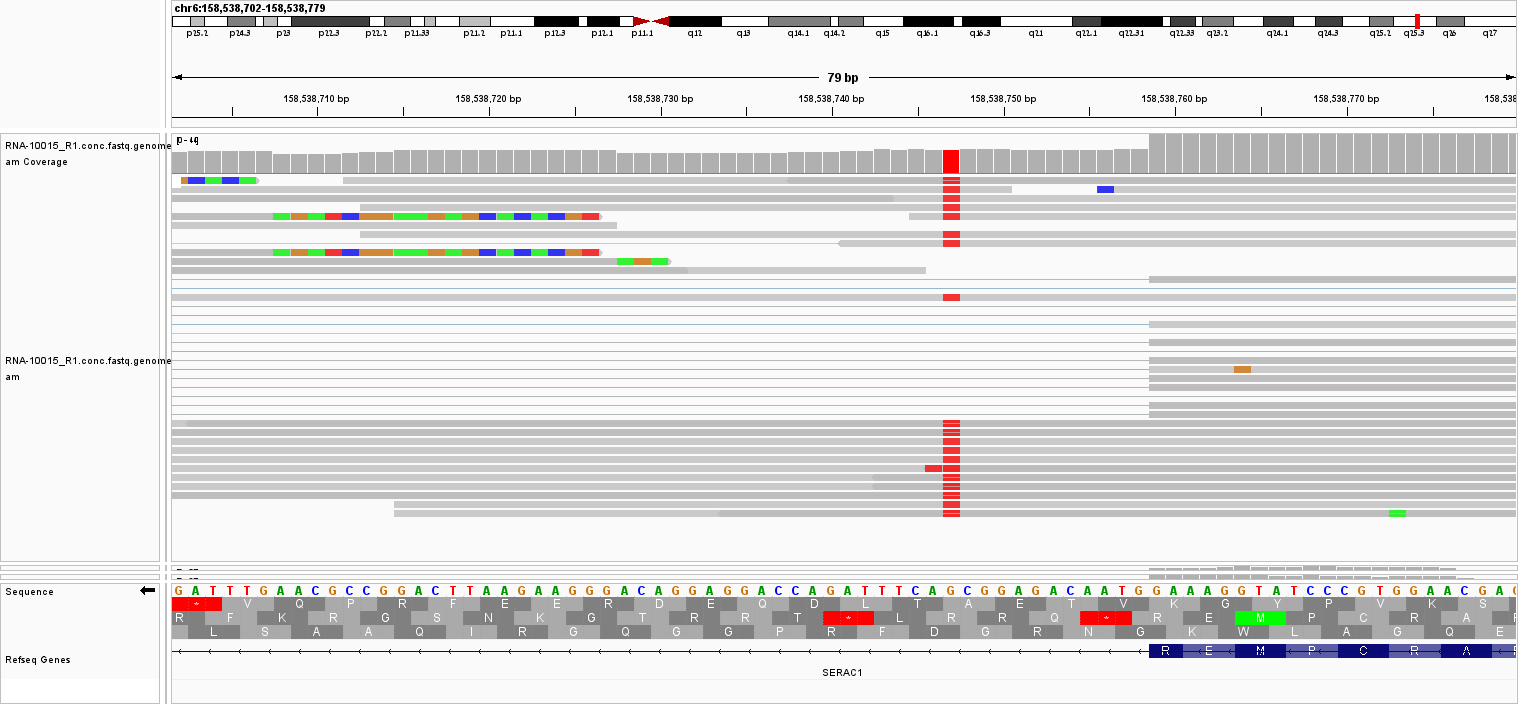

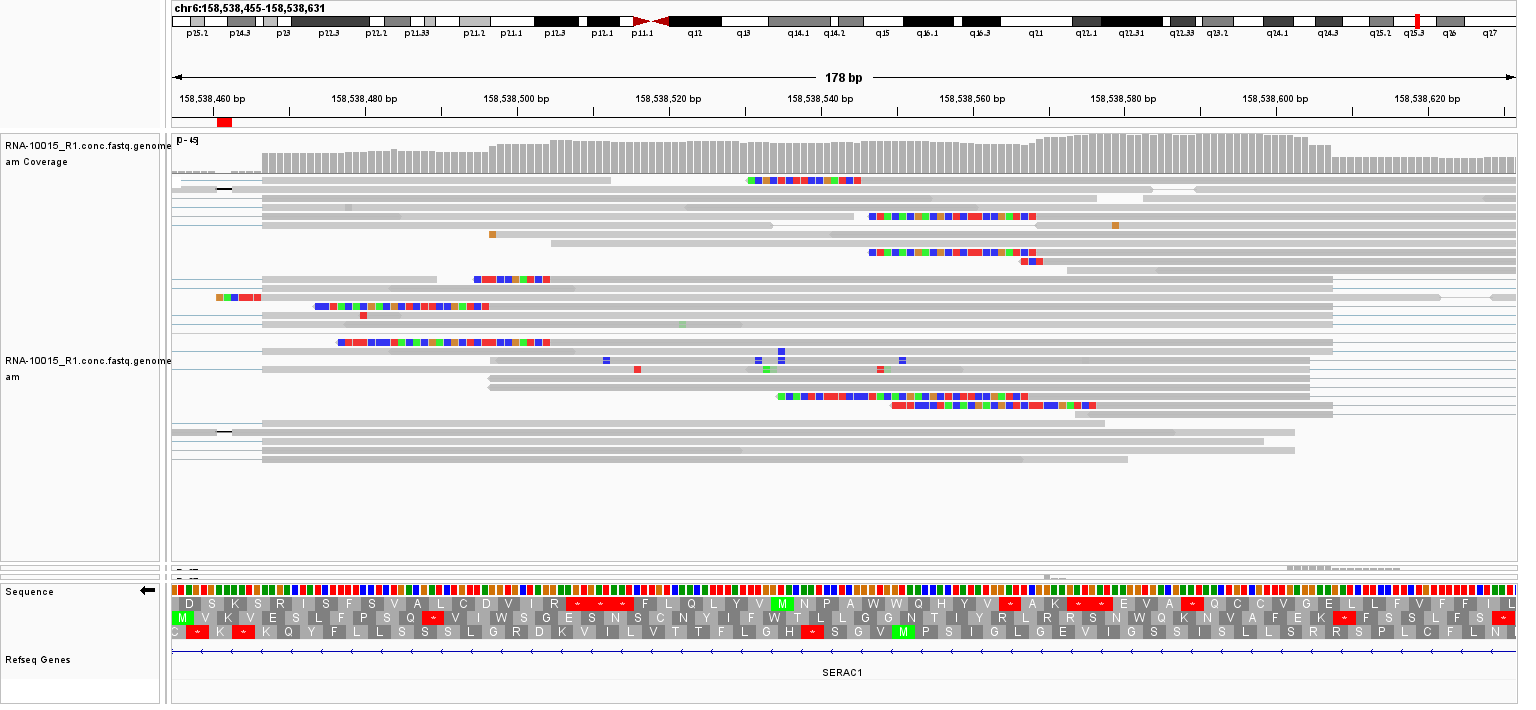

**Figure S6: Functional Consequences of the Intron Inclusion and Retention in *SERAC1* 1403+297_1403+298 delAA variant.**
Upper panel: Schematic representation of the alternative splicing outcome caused by the variant, leading to an aberrant intronic retention and premature termination of translation.
Lower panel: In another transcript, the pseudo-exon activation results in a frameshift and premature stop codon which may indicate a loss-of-function mechanism.

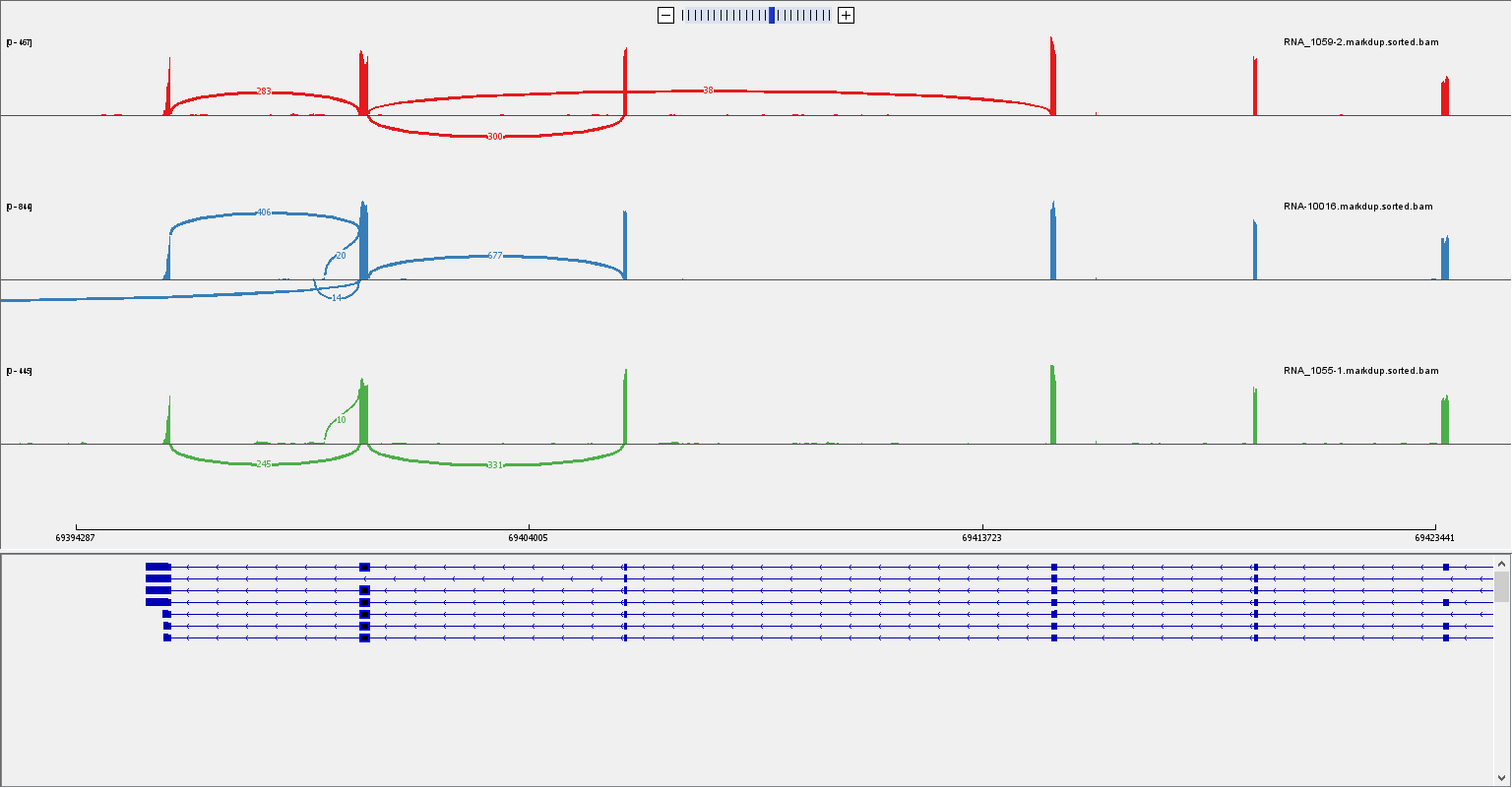

Exon 6

Control 2

Control 1

Affected

**Figure S7: Aberrant splicing associated with the *NFU1*:c.545G>A / p.Arg182Gln paternally inherited variant- first transcript.**
Sashimi plot illustrating the surrounding junctions of exon 6, of *NFU1* in the affected individual and 2 controls. The red arrow points exon 6 skipping in the affected sample, which is demonstrated in 38 reads vs. 300 wild-type reads (~11%). Genomic positions in GRCh38.

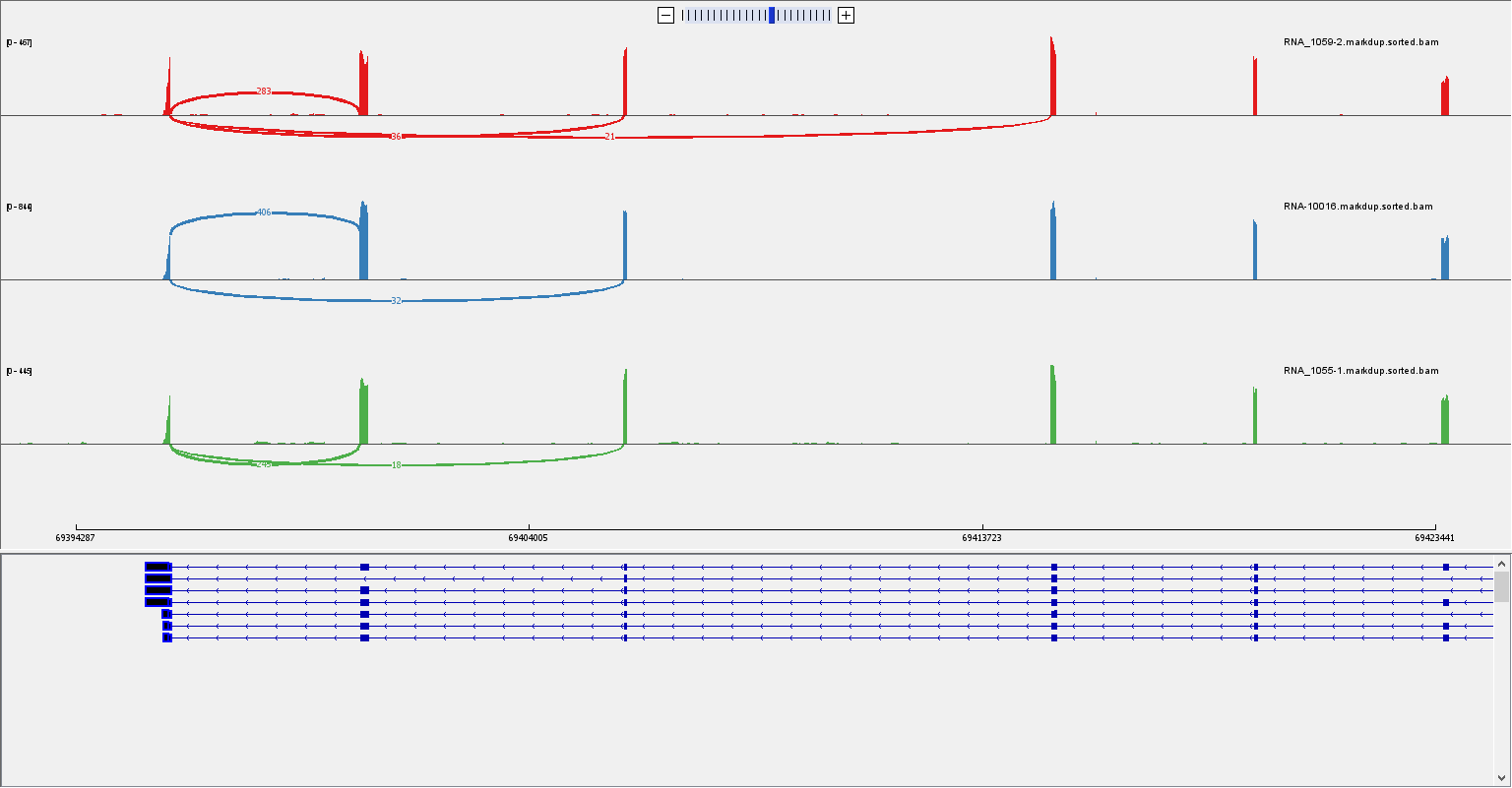

Exon 7

Exon 6

Control 2

Control 1

Affected

**Figure S8: Aberrant splicing associated with the *NFU1*:c.545G>A / p.Arg182Gln paternally inherited variant- second transcript.**
Sashimi plot illustrating the surrounding junctions of exon 6 and 7 of *NFU1* in the affected individual and 2 controls. The red arrow points exons 6-7 skipping which was demonstrated in 21 reads (~6% of all reads). Genomic positions in GRCh38.
